# Development of reference equations for the six-minute walk distance of school-aged Nigerian children

**DOI:** 10.1101/2021.12.21.21267745

**Authors:** Peter Odion Ubuane, Olufunke Adewumi Ajiboye, Motunrayo Oluwabukola Adekunle, Ayodeji Olushola Akinola, Gbenga Akinyosoye, Mogbafolu Olugbemiga Kayode-Awe, Omotola Aderiyike Ajayi, Chidimma Imma Ohagwu, Barakat Adeola Animasahun, Fidelis Olisamedua Njokanma

## Abstract

**Background:** The **six-minute walk test** (*6MWT*), a simple, reliable and valid test that uses the distance walked in six minutes (six-minute walk distance, *6MWD*) to quantify *functional capacity*, is widely used in chronic cardiopulmonary and non-cardiopulmonary disorders. However, absence of reference standards for Nigerian school-age children limits its use in this age group.

**Objectives:** To develop normative values and equations for the 6MWT of school-aged Nigerian children.

**Methods:** In a cross-sectional study, healthy children aged 6-11 years in Lagos, Nigeria, completed 6MWT on 20-meter straight outdoor courses in their schools following standardized guidelines. *Potential predictors*: demographic (age, sex), anthropometric (height, weight, chest circumference, leg length) and physiologic data [pre-walk, immediate post-walk and 5^th^ minute-post-walk heart rate (*HR*), oxygen saturation (*SpO*_*2*_), systolic pressure (*SBP*), diastolic blood pressure (*DBP*) and rating of perceived exertion (*RPE*)] and difference between pre-walk and post-walk HR (Δ*HR)*, SpO_2_ ((Δ*SpO*_*2*_), SBP (Δ*SBP)*, DBP (Δ*DBP*) and RPE (Δ*RPE*). *Primary outcome*: six-minute walk distance (6MWD) in metres.

**Results:** Overall, 627 pupils (52.1% girls) walked 504.6 ± 66.6 m (95% CI: 499.4, 509.8), ranging from 326.6 m to 673.0 m; 16 m longer in boys (p=0.002). Stepwise linear regression yielded: 6MWD (m) = 347.9 + 14 (Age) + 1.6 (ΔHR) + 17.6 (Sex) + 1.2 (ΔSBP); *R*^*2*^ =0.25. Previously published reference equations over-estimated Nigerian children’s 6MWD.

**Conclusion:** The reference values and prediction equations, after validation in other Nigerian populations, may be useful for functional evaluation of Nigerian children aged 6-11 years with chronic childhood disorders.

## INTRODUCTION

Functional exercise capacity, which is the ability to conduct day-to-day activities, is frequently impaired in chronic cardio-pulmonary and non-cardiopulmonary disorders. This functional impairment worsens quality of life, morbidity and mortality, independent of the effect of the underlying disease.(1–3) Thus, detecting, quantifying and managing this functional impairment is required for optimal management of chronic disorders.(4, 5) The *gold-standard* objective measure of functional capacity-the maximal oxygen consumption (*VO*_*2max*_) from a cardiopulmonary exercise test (CPET)-(6) requires sophisticated equipment and expertise largely unavailable in developing countries.(4, 7, 8) But, the six-minute walk test (6MWT), which evolved from pre-existing field walk tests, (1, 9, 10) is a cheap but valid and reliable potential alternative measure of functional capacity using the self-paced distance walked in six minutes (six-minute walk distance, 6MWT) on a smooth hard surface. It is widely used for the functional evaluation, prognostication and rehabilitation of pediatric and adult populations with cardiopulmonary and cardiopulmonary disorders (4, 7, 10–13)

However, its clinical utility in sub-Saharan Africa is limited largely because of lack of locally-derived normative standards.(14–17) Published references from other ethno-geographic populations are unreliable when applied to another because of wide variations in the mean 6MWD and its predictors among studies even from same country.(1, 7, 16, 18) Although, the 6MWD of children is mostly influenced by age and height, the effect of other factors like sex, weight or body mass index (BMI) vary widely among studies.(14–16, 18–20) The American Thoracic Society (ATS) and European Respiratory Society (ERS) thus recommend development of reference standards from local populations.(7) In Nigeria, although reference standards are available for adolescents(21) and adults(8, 22), none is available in pre-adolescents school-age children. This cross-sectional study thus determined the mean 6MWD of healthy 6 to 11-year-old Nigerian children, its predictors (including chest circumference, which has not been previously evaluated) and reference equations; we also assessed the predictive performance of previously published equations. We used 20 m long tracks, rather than ATS-recommended 30m long tracks for adults, because shorter tracks are more likely to be available in schools and hospitals.(23, 24). Parts of these data have been published as a dissertation and conference abstract.(25)

## METHODS

Our study design, sample selection, data collection/measurements and statistical analyses procedures are detailed in the online supplement.

### POPULATION

In this cross-sectional study, we used multi-stage random sampling to select Nigerian children aged 6-11 years from primary schools within Ikeja Local Government Area of Lagos State, Nigeria, excluding those with cardiopulmonary, hematologic, neuro-musculoskeletal disorders and those with recent respiratory infections or hospitalization.

### SAMPLE SIZE

Although we needed at least 417 pupils to estimate the mean 6MWD of Nigerian children within 1% precision (sample size formula for single mean, based on standard deviation of 73 m among North African children(26)), we exceeded this minimum size to enhance precision of our statistical estimates and prediction equations.(27)

### ETHICS

The study was approved by the Health Research & Ethics Committee of the Lagos State University Teaching Hospital (LREC/10/06/486), Lagos State Universal Basic Education and Ministry of Education. We also obtained written parental consent and pupil’s assent.

### DATA COLLECTION

We conducted field work from April 2016 to June 2017 (8.00AM - 2.30PM).

#### Baseline pre-walk measurements and instructions

Before the 6MWT, each participant rested for 5-10 minutes, during which we conducted standardized anthropometric (weight, height, leg length and chest circumference), spirometric (peak expiratory flow, *PEF*; forced expiratory volume in 1 second, *FEV*_*1*_), blood pressure (systolic blood pressure, *preSBP*; diastolic blood pressure, *preDBP*), oximetric (oxygen saturation, *preSpO*_*2*_); heart rate (*preHR*) and rating of perceived exertion (*preRPE*) measurements.

#### 6MWT procedure

We followed ATS/ERS standardized test instructions to administer the 6MWT on one participant at a time on 20-m long straight tracks on outdoor flat, smooth-surfaced corridors or paved walkways in each school.(1, 5, 7) We instructed participants to walk as *far as they can walk* without running/jogging; they could rest or stop during the test while we continued timing. At the end of six minutes, we measured the distance walked (6MWD) and repeated the physiological measurements immediately (*postHR, postSpO*_*2*_, *postSBP, postDBP, postRPE*) and after a five-minute rest (*5mHR, 5mSpO*_*2*_, *5mSBP, 5mDBP*). We also asked for specific symptoms felt while walking.

### DATA MANAGEMENT

We conducted statistical analyses with Bluesky® Statistics version 7.30 and JASP® statistics version 0.14 (graphical user interphases for R) with significance at p value < 0.05 at 95% confidence interval (CI). We computed: change in physiological measurements (Δ*HR*, Δ*SpO*_*2*_, Δ*SBP*, Δ*DBP*, Δ*RPE)* as the difference between the corresponding immediate post-walk and baseline values; predicted maximal HR as (*PredHR*_*max*_)= 208 – (0.7 x age in years)(28); and percentage predicted maximal heart rate (%*PredHR*_*max*_) as (*postHR*/*PredHR*_*max*_) x100%. We conducted univariable (descriptives), bivariable (student’s t-test, Mann-Whitney U-test, paired t-test, Wilcoxon signed-rank test, ANOVA, linear correlation, chi square, Bland-Altman analysis) and multivariable analyses (stepwise multivariable linear regression). The effect sizes of statistically significant comparisons were quantified with Cohen’s d (trivial: < 0.1, small/weak: 0.2, medium/moderate: 0.5 and large/strong: 0.8), rank biserial or Pearson’s correlation (trivial: < 0.1, small: 0.1, medium/moderate: 0.3 and large/strong: 0.5) and partial eta squared, *η*^*2*^*p* (trivial: < 0.1, small: 0.01, medium/moderate: 0.06 and large/strong: 0.14).

## RESULTS

We recruited 956 pupils from 14 schools (9 public schools). Of this, 627 completed the 6MWT (Figure 1).

**Figure 1:**
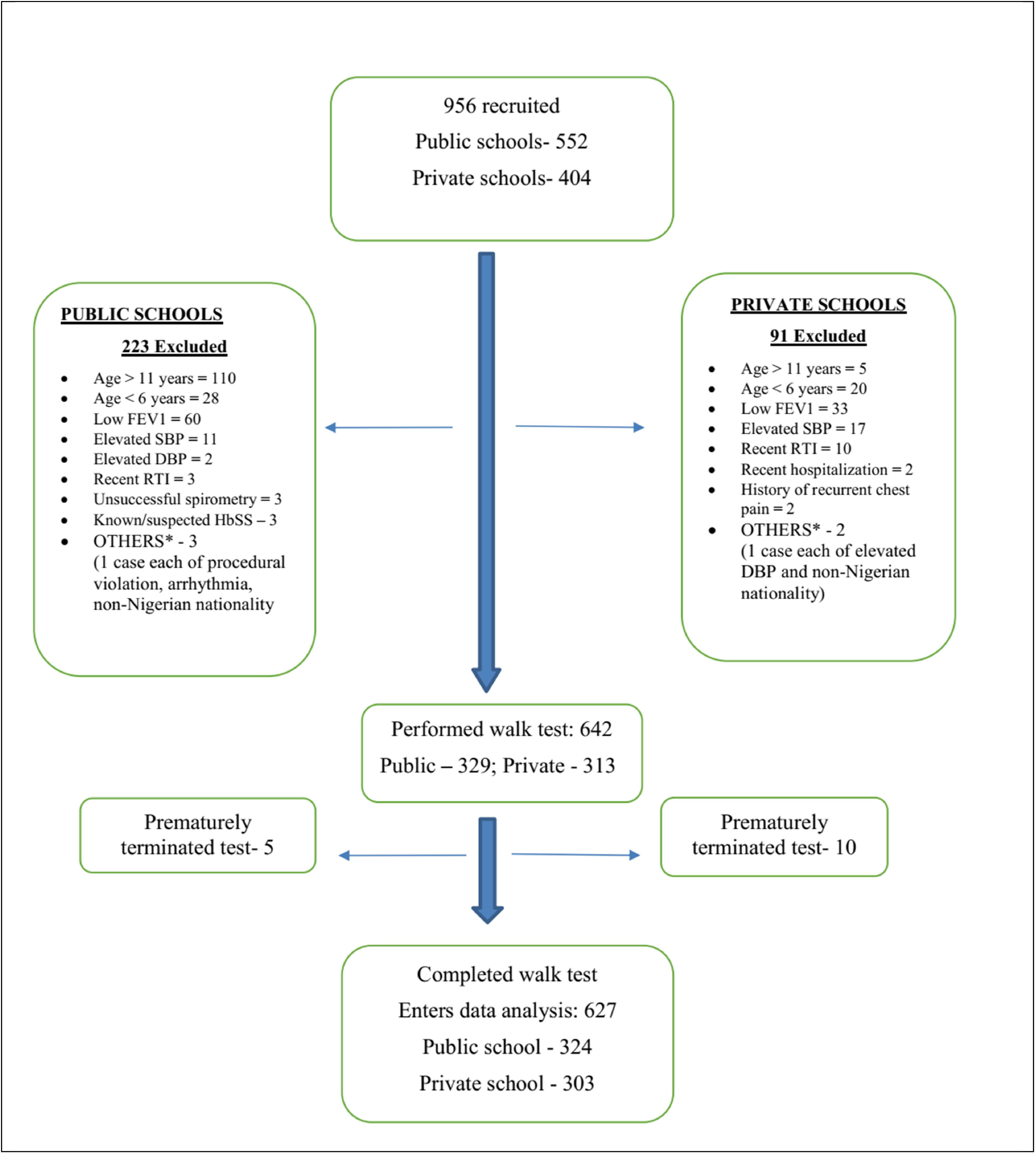
Participants’ Flow chart. DBP: diastolic blood pressure; SBP: systolic blood pressure; RTI: respiratory tract infection; low FEV1 was defined as FEV1 < 80% of predicted local reference; high DBP and SBP were defined as > 90th percentile using CDC’s chart

### DEMOGRAPHIC, ANTHROPOMETRIC AND NUTRITIONAL CHARACTERISTICS

About half (52.1%) were girls, almost half (42 %) were of Yoruba ethnicity and over one-quarter (28.7%) were overweight/obese (Table 1). Overall, girls and boys had comparable age, weight, height, BMI, chest circumference, leg length and school type and nutritional status (p > 0.05 for all variables). However, 11-yr old girls were taller than their male counterparts (Table E2).

**Table 1.**
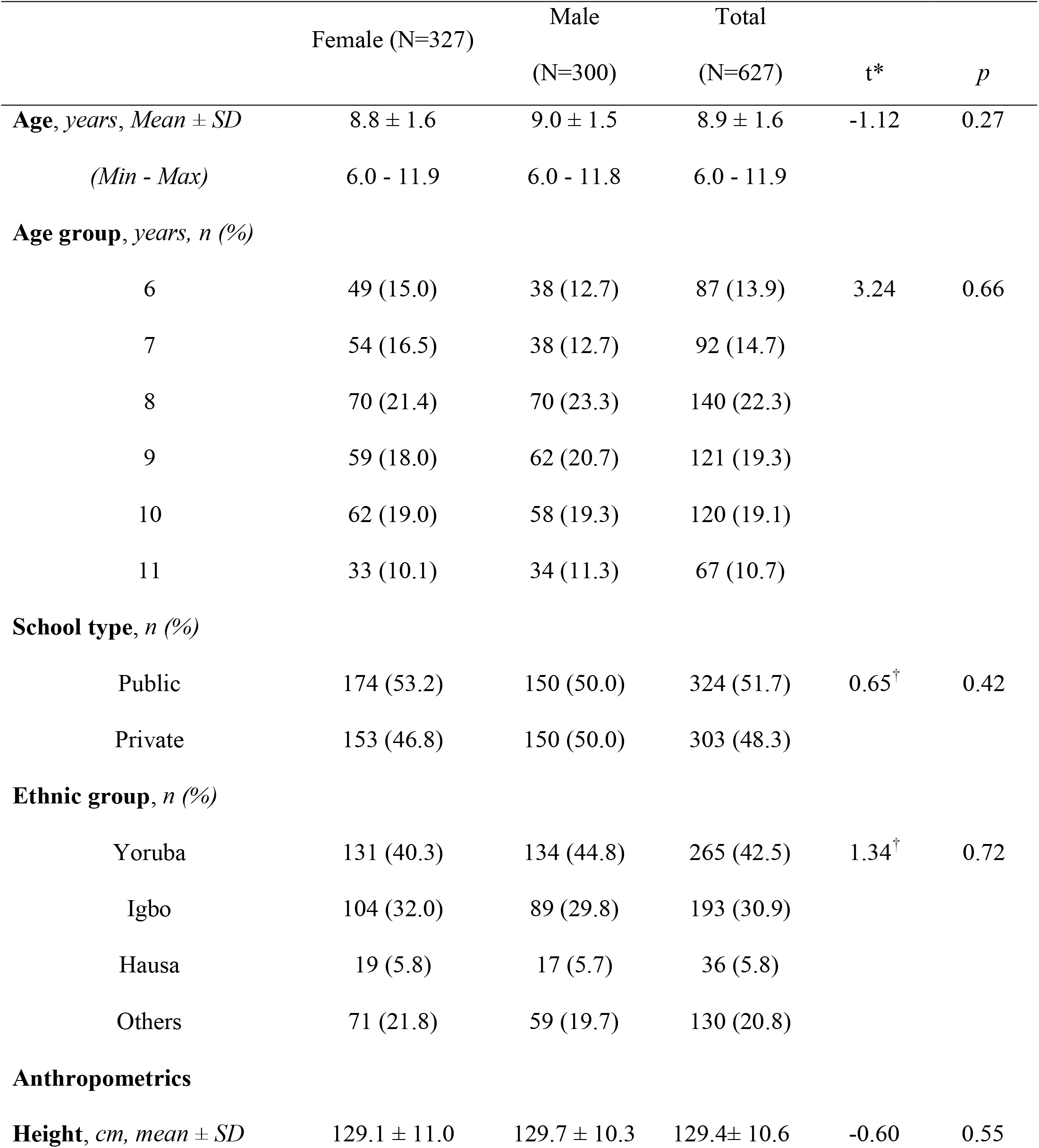
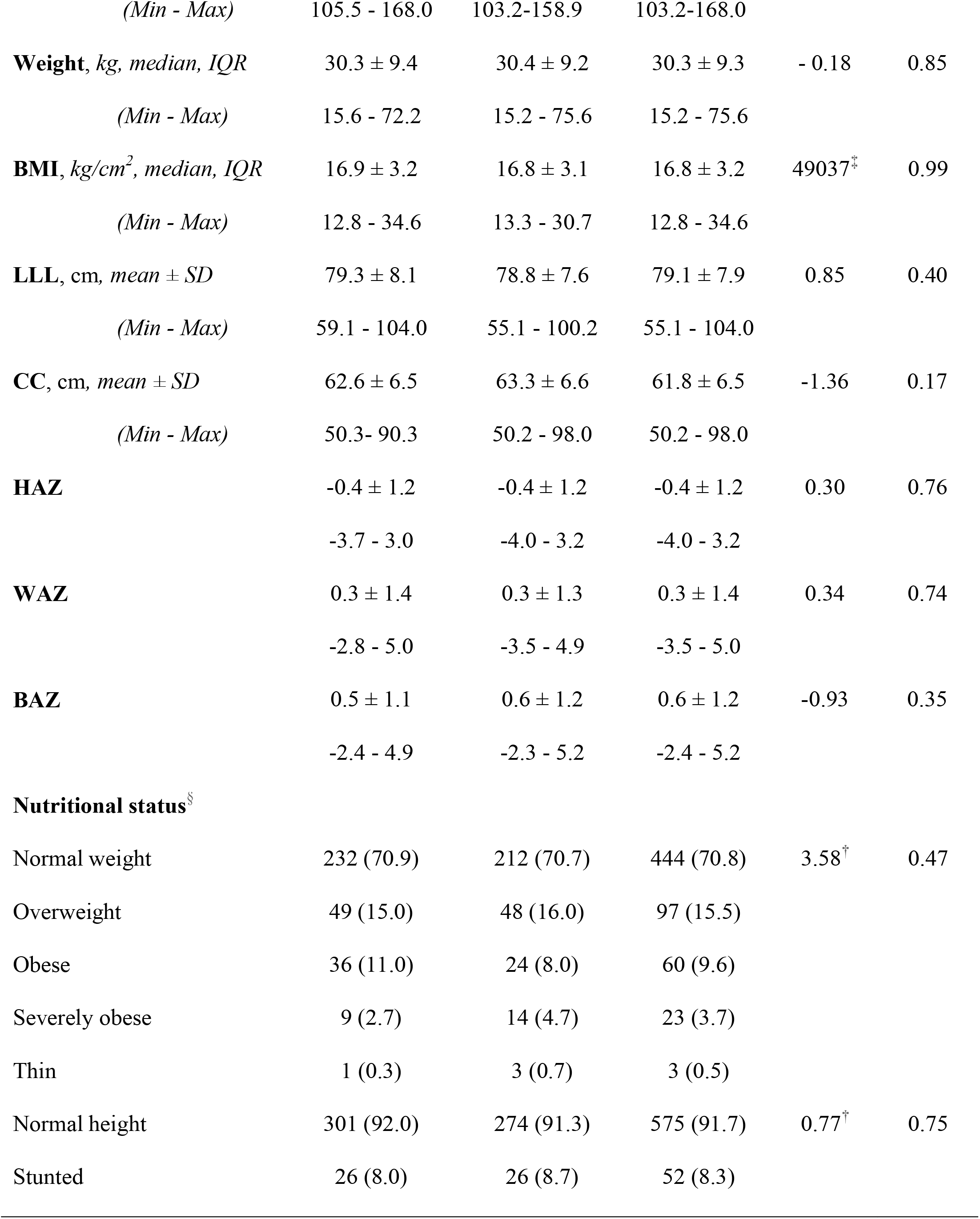
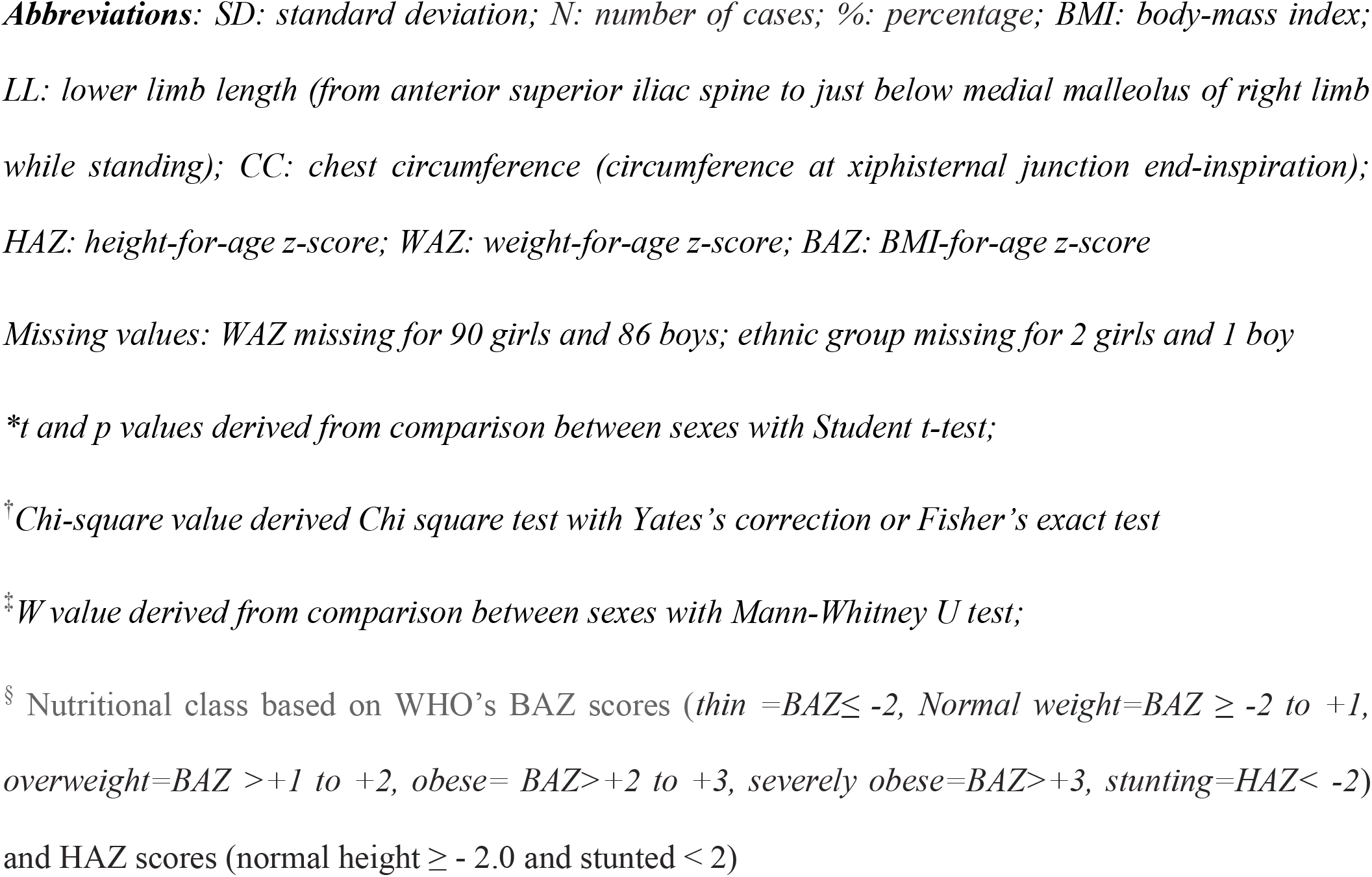
Demographic, anthropometric and nutritional characteristics of study participants

### SIX-MINUTE WALK DISTANCE

Overall, 6MWD ranged from 326.6 m to 673.0 m, with mean ± SD of 504.6 ± 66.6 m (95% CI: 499.4, 509.8) (Table 2).

**Table 2.**
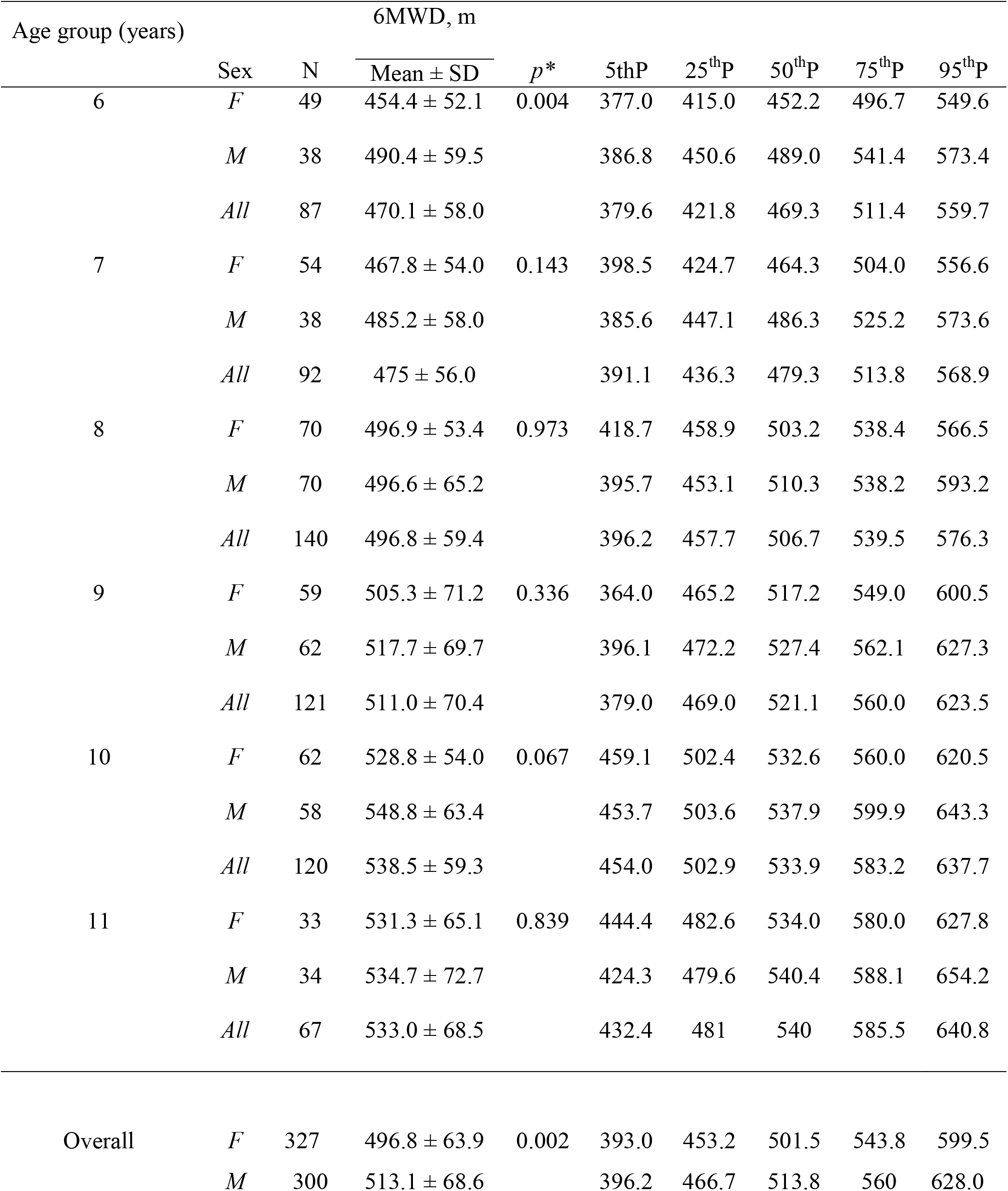
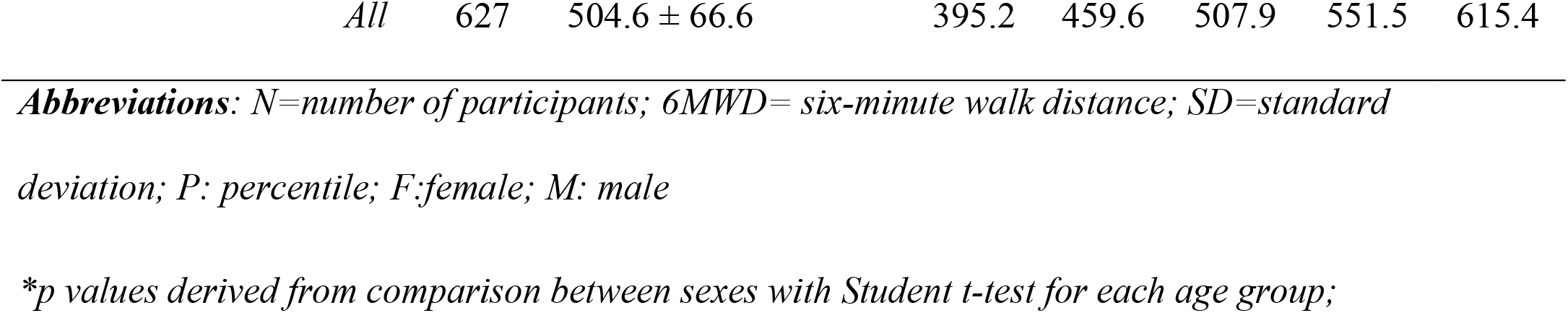
Mean and percentile values for the six-minute walk distance of Nigerian children

### PREDICTORS OF THE 6MWD OF NIGERIAN CHILDREN

#### Sex and age

Boys walked 16.3 m longer than girls [513.1 (95% CI: 505.3, 520.9) vs 496.8 (95% CI: 489.8, 503.7); p = 0.002; Cohen’s d (95% CI) = -0.25 (−0.40, -0.09)]. The 6MWD increased linearly by 62.9 m between ages six to eleven years [F (5,621) = 20.31, p < 0.001, η^2^p = 0.14, linear trend p < 0.001; Fig 2), with the steepest year-to-year increase of 26.8 m between 9 to 10 years (t = -3.35, p = 0.006). In each age-group, there was no significant sex difference in 6MWD except amongst six-year olds (p = 0.004) (Table 2). Between ages 10 and 11, the walk distance plateaued in girls and dipped in boys and the total cohort (Fig 2).

**Fig 2:**
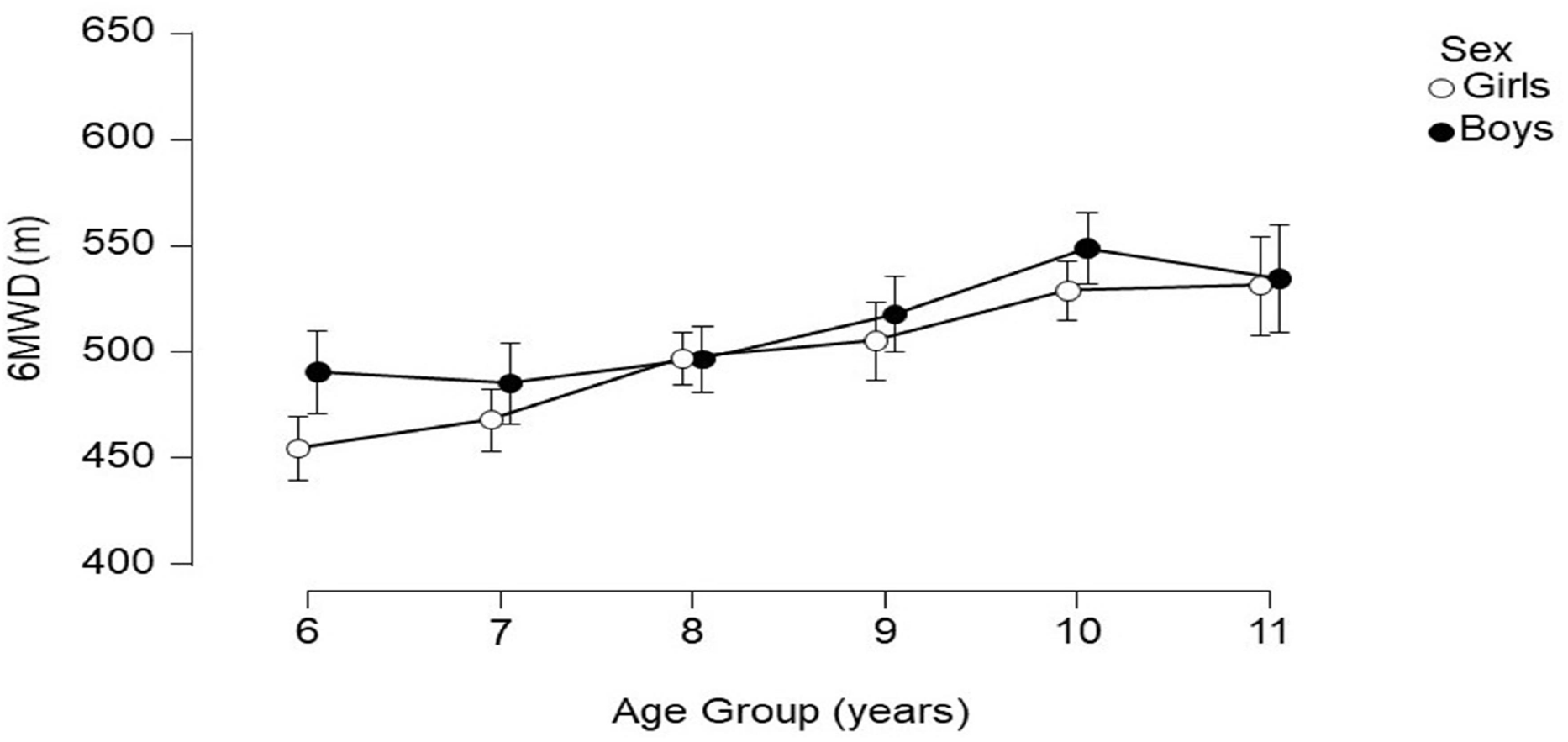
Change in Six-minute walk distance with increasing age in girls and boys Abbreviations: 6MWD, six-minute walk distance in metres;

#### BIVARIABLE LINEAR CORRELATION

Overall, 6MWD moderately positively correlated with age (r=0.37), height (r=0.32), PEF (r=0.31) and *postSBP* (0.30); moderately negatively correlated with *PredHR*_*max*_ (r=-0.37); and weakly positively correlated with *%*Δ*HR* (r=0.29), FEV1 (r=0.29), Δ*HR* (r=0.28), LLL (r=0.28), Δ*SBP* (r=0.27), weight (r=0.19), *%PredHR*_*max*_ (r=0.12) and *preRPE* (r=0.10) (Table 3).

**Table 3.** Bivariable linear correlation between 6MWD and continuous variables

| Variables | Girls |  |  | Boys |  |  | All |  |  |
| --- | --- | --- | --- | --- | --- | --- | --- | --- | --- |
|  | *n* | *r* | *p* | *n* | *r* | *p* | *n* | *r* | *p* |
| Age | 327 | 0.41 | <0.0001 | 300 | 0.32 | <0.0001 | 627 | 0.37 | <0.0001 |
| *PredHR<sub>max</sub>* | 327 | -0.41 | <0.0001 | 300 | -0.32 | <0.0001 | 627 | -0.37 | <0.0001 |
| Height | 327 | 0.38 | <0.0001 | 300 | 0.25 | 0.0035 | 627 | 0.32 | <0.0001 |
| *PEF* | 327 | 0.34 | <0.0001 | 300 | 0.26 | 0.001 | 627 | 0.31 | <0.0001 |
| *PostSBP* | 327 | 0.30 | <0.0001 | 300 | 0.31 | <0.0001 | 627 | 0.30 | <0.0001 |
| *%ΔHR* | 326 | 0.26 | <0.0001 | 299 | 0.34 | <0.0001 | 625 | 0.29 | <0.0001 |
| *FEV<sub>1</sub>* | 327 | 0.34 | <0.0001 | 300 | 0.21 | 0.0655 | 627 | 0.29 | <0.0001 |
| *ΔHR* | 326 | 0.25 | 0.001 | 299 | 0.34 | <0.0001 | 625 | 0.28 | <0.0001 |
| LLL | 327 | 0.34 | <0.0001 | 300 | 0.23 | 0.019 | 627 | 0.28 | <0.0001 |
| *ΔSBP* | 327 | 0.25 | 0.002 | 300 | 0.31 | <0.0001 | 627 | 0.27 | <0.0001 |
| Weight | 327 | 0.25 | <0.0001 | 300 | 0.13 | 0.05 | 627 | 0.19 | <0.0001 |
| *%PredMaxHR* | 327 | 0.07 | 0.702 | 300 | 0.25 | <0.0001 | 627 | 0.12 | 0.005 |
| *PreRPE* | 320 | 0.06 | 0.431 | 298 | 0.15 | 0.05 | 618 | 0.10 | 0.035 |
| *PostHR* | 327 | 0.05 | 1.000 | 300 | 0.23 | 0.011 | 627 | 0.11 | 1.000 |
| *PostRPE* | 324 | -0.07 | 0.431 | 294 | 0.12 | 0.08 | 618 | 0.02 | 1.000 |
| BMI | 327 | 0.06 | 0.254 | 300 | 0.0012 | 0.98 | 627 | 0.03 | 0.389 |
| $\Delta RPE$ | 317 | -0.1 | 0.272 | 292 | 0.07 | 0.21 | 609 | -0.02 | 1.000 |
| $PreHR$ | 326 | -0.17 | 0.4249 | 299 | -0.03 | 1.000 | 625 | -0.12 | 0.42 |

**Abbreviations:** $n$ = number; $r$ = Pearson's correlation coefficient; $*p$ values are adjusted for multiple comparisons with Bonferoni-Holms' test; $PredHR_{max}$ = predicted maximal heart rate in beats per minute; $PEF$ =peak expiratory flow in litres per minute; $PostSBP$ : immediate post-walk systolic blood pressure; $\% \Delta HR$ = percentage of heart rate change from baseline to immediate post-walk; $FEV_1$ = forced expiratory volume in 1 second in litres; $\Delta HR$ = heart rate change from baseline to immediate post-walk; $LLL$ = lower limb length in cm; $\Delta SBP$ = change in SBP from baseline to immediate post-walk in mmHg; $\%PredMaxHR$ = percentage of predicted maximal heart rate;
$PreRPE$ : pre-walk/baseline rating of perceived exertion; $PostHR$ : immediate post-walk HR; $PostRPE$ : immediate post-walk rating of perceived exertion; $BMI$ = body-mass index in $kg/cm^2$ ; $\Delta RPE$ = change in rating of perceived exertion from baseline/pre-walk to immediate post-walk.
For each of the following variables not shown in this table, $r$ was $\leq 0.14$ and $p$ was 1.000 in both sexes and in the total sample: $PreSBP$ (baseline SBP), $PreDBP$ (baseline DBP), $PostDBP$ (immediate post-walk SBP) , chest circumference, $PreSpO_2$ (baseline oxygen saturation), $PredPEF$ (percentage predicted PEF), $HAZ$ (height for age z-score), $PostSpO_2$ (immediate post-walk $SpO_2$ ), $\Delta DBP$ (change in DBP from baseline to immediate post-walk DBP), $\Delta SpO_2$ (change in $SpO_2$ from baseline to immediate post-walk $SpO_2$ ), $PredFEV_1$ (percentage predicted $FEV_1$ ), $WAZ$ (weight for age z-score) and $BAZ$ (BMI for age z-score)

#### OTHER POTENTIALLY CONFOUNDING FACTORS

Table 4 shows that pupils in public schools walked 18.8 m longer distance than those in private schools [p < 0.0001, Cohen’s d = -0.28 (−0.44, -0.13)]. However, 6MWD was not influenced by ethnicity (p = 0.244), time of testing (p = 0.205), overweight/obesity (p = 0.077), stunting (p = 0.288) or participation in extra-curricular sports (p = 0.270). Compared to pupils in public schools, those in primary schools were significantly taller, heavier, and had wider chest circumference and longer lower limb length, despite similar mean age.

**Table 4.**
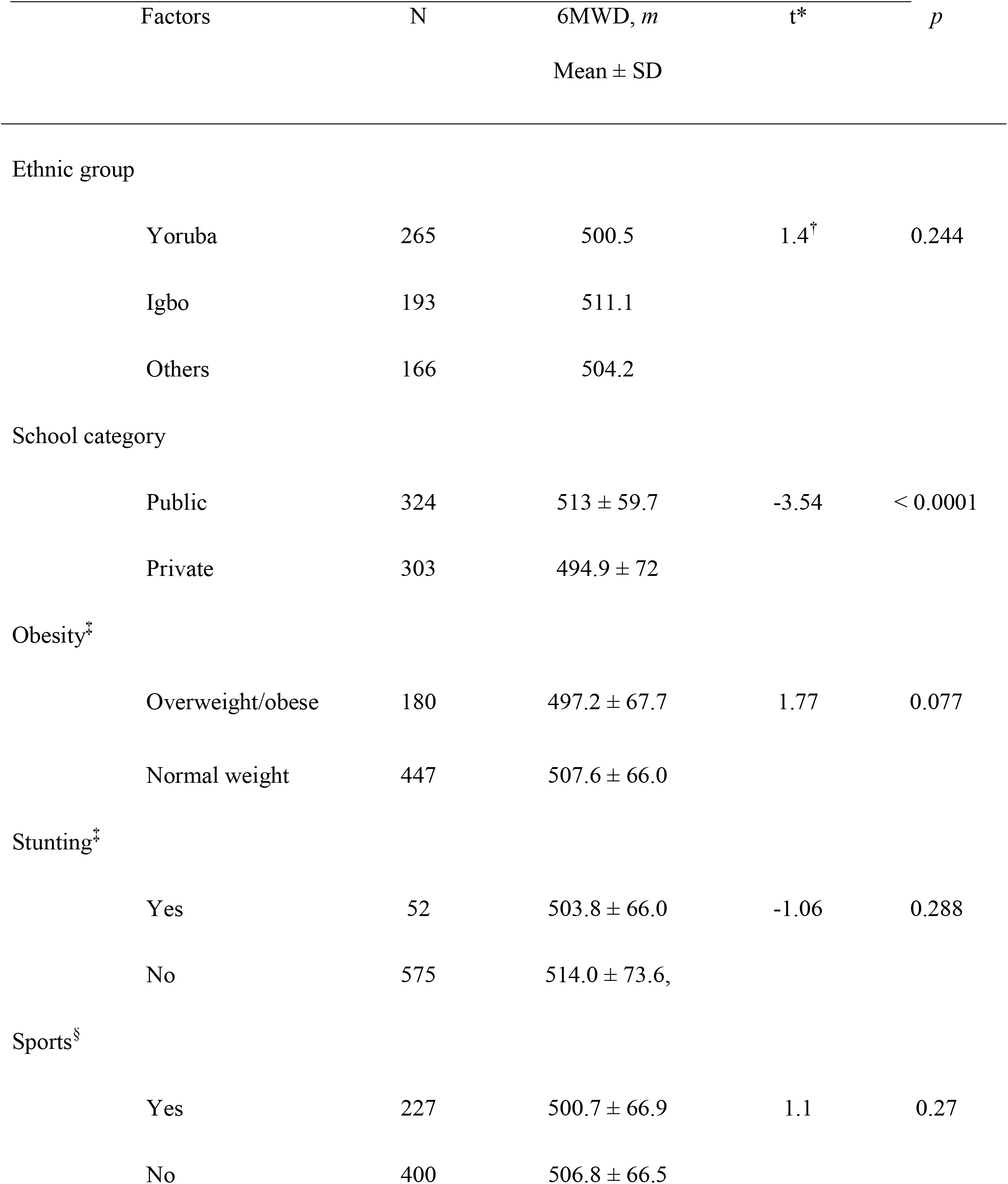
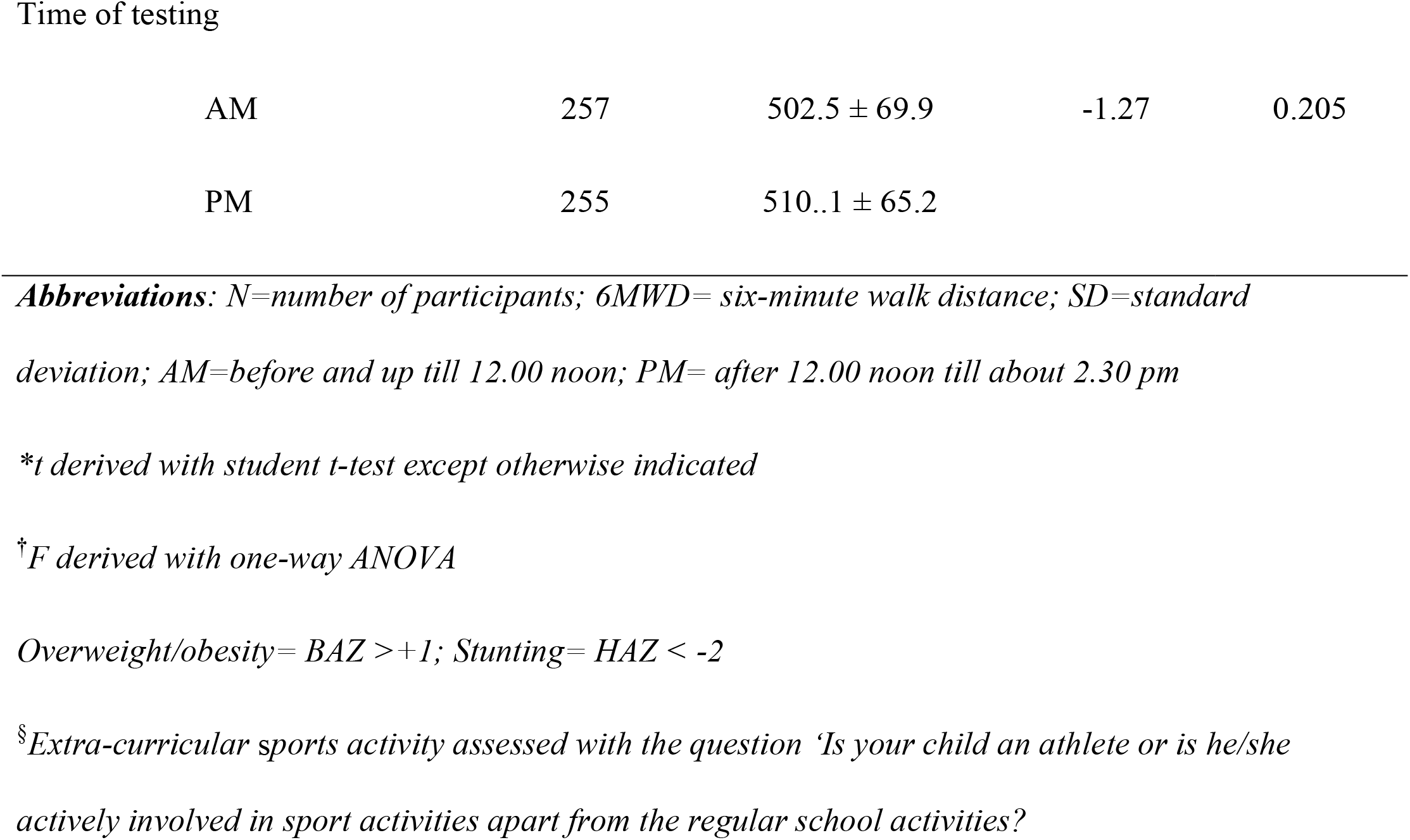
Other potentially confounder variables for the 6MWD

| Factors | N | 6MWD, *m*<br>Mean ± SD | *t* * | *p* |
| --- | --- | --- | --- | --- |
| Ethnic group |  |  |  |  |
| Yoruba | 265 | 500.5 | 1.4<sup>†</sup> | 0.244 |
| Igbo | 193 | 511.1 |  |  |
| Others | 166 | 504.2 |  |  |
| School category |  |  |  |  |
| Public | 324 | 513 ± 59.7 | -3.54 | < 0.0001 |
| Private | 303 | 494.9 ± 72 | | |
| Obesity<sup>‡</sup> |  |  |  |  |
| Overweight/obese | 180 | 497.2 ± 67.7 | 1.77 | 0.077 |
| Normal weight | 447 | 507.6 ± 66.0 | | |
| Stunting<sup>‡</sup> |  |  |  |  |
| Yes | 52 | 503.8 ± 66.0 | -1.06 | 0.288 |
| No | 575 | 514.0 ± 73.6, | | |
| Sports<sup>§</sup> |  |  |  |  |
| Yes | 227 | 500.7 ± 66.9 | 1.1 | 0.27 |
| No | 400 | 506.8 ± 66.5 | | |
| Time of testing |  |  |  |  |
| AM | 257 | 502.5 ± 69.9 | -1.27 | 0.205 |
| PM | 255 | 510.1 ± 65.2 |  |  |

**Abbreviations:** *N*=number of participants; 6MWD= six-minute walk distance; SD=standard deviation; AM=before and up till 12.00 noon; PM= after 12.00 noon till about 2.30 pm
*\*t derived with student t-test except otherwise indicated*
*†F derived with one-way ANOVA*
*Overweight/obesity= BAZ >+1; Stunting= HAZ < -2*
*§Extra-curricular sports activity assessed with the question 'Is your child an athlete or is he/she actively involved in sport activities apart from the regular school activities?'*

#### PHYSIOLOGICAL RESPONSES DURING THE 6MWT

Changes in heart rate, SpO_2_, BP and RPE are detailed and illustrated (Figure E2, Table E3) in the supplementary file.

#### POST-WALK SYMPTOMS

About one-third (190/627, 30.3%) of our participants had one or more symptoms when specifically asked at the end of the walk test, the commonest being leg pain (19.6%) (Table 5). The symptoms were mild and resolved at the stop of the walk test. None of this group terminated the test while walking.

**Table 5.** Symptoms experienced by healthy Nigerian children during 6MWT

|  | Sex |  | Total |
| --- | --- | --- | --- |
|  | Girls | Boys | N (%) |
| Exercise-induced symptoms | N (%) | N (%) |  |
| Chest Pain | 28 (8.6) | 25 (8.3) | 53 (8.4) |
| Chest Tightness | 18 (5.5) | 17 (5.7) | 35 (5.6) |
| Dizziness | 21 (6.4) | 23 (7.7) | 44 (7.0) |
| Palpitation | 17 (5.2) | 12 (4.0) | 29 (4.6) |
| Leg Cramps | 0 | 2 (0.7) | 2 (0.3) |
| Leg Pains | 59 (18.0) | 64 (21.3) | 123 (19.6) |
| Total* | 97 (29.7) | 93 (31.1) | 190*(30.3) |

*Abbreviations: N= number of participants; %= percentage of total number of girls (n=327), total number of boys (n=300) and total cohort (n=627)*
*\*Total counts exceeds 97 because some children had more than one symptom*

#### MULTIVARIABLE ANALYSIS

An initial exploratory stepwise linear regression model (Table E4) was further simplified to yield 6MWD (m) = 347.9 + 14*Age* + 1.6Δ*HR* + 17.6*Sex* + 1.2ΔS*BP* (*R*^*2*^*=0*.*25*) and two additional equations which may serve as alternative when blood pressure and heart rate are not measured as part of a 6MWT (Table 6). All the three models satisfied the assumptions of multiple linear regression (Table E5 and Figure E3).

**Table 6:**
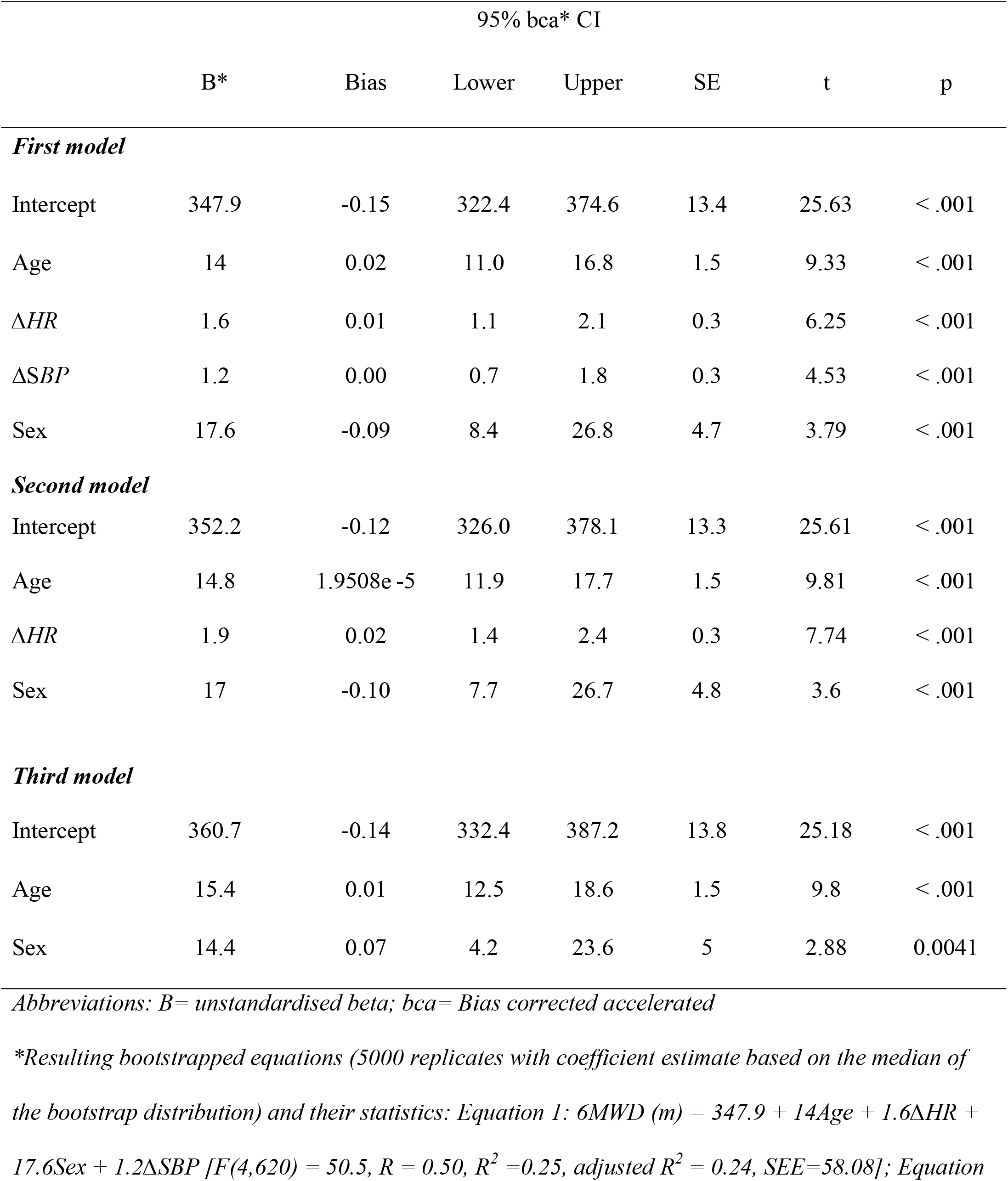
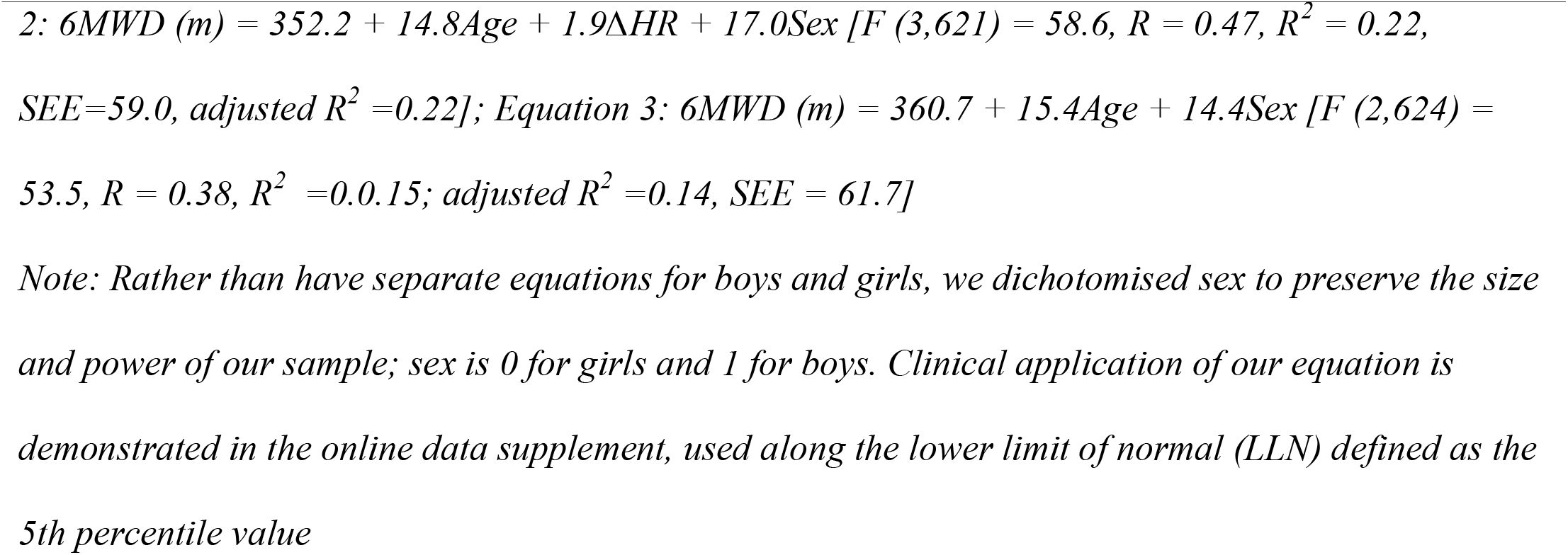
Bootstrapped regression models for the 6MWD of Nigerian children aged 6-11 years

#### Comparison with previously published reference equations

Although the Taiwanese equation by Chen et al(29) correlated excellently with our measured 6MWD, with small bias and acceptable LoA in Bland-Altman analysis (Table 7), the plot showed a proportional bias, tending towards over-estimation with increasing age (Figure 3C). Equations from Nigerian adolescents, Brazilian and Swiss children correlated moderately with our 6MWD with small biases, however the LoAs were wide (Figure 3A,J,K). Other equations over-estimated the 6MWD of Nigerian children with wide limits of agreement (LoA) despite weak to moderate correlations.

**Table 7.** Comparison of the 6MWD of Nigerian children with previously published regression equations.

| First Author | Country | Track | Age range<br>(years) | *r* | *p* | *d*<br>(lower, upper) |
| --- | --- | --- | --- | --- | --- | --- |
| Geiger | Austria | 30 m | 3 -18 | 0.38 | <0.0001 | -103.9<br>(-227.0, 19.2) |
| Garcia | Venezuela | 30 m | 6 - 11 | 0.37 | <0.0001 | -188.0<br>(-315.1, -60.9) |
| Ulrich | Switzerland | 30 m | 5 - 17 | 0.31 | <0.0001 | -25.6<br>(-150.1, 99.0) |
| Priesnitz | Brazil | 30 m | 6 - 12 | 0.45 | <0.0001 | -45.9<br>(-162.6, 70.8) |
| Oliveira | Brazil | 22 m | 6 - 13 | 0.40 | <0.0001 | -194.9<br>(-316.7, -73.1) |
| Saad | Tunisia | 40 m | 6 - 16 | 0.40 | <0.0001 | -250.6<br>(-267.6, -141.3) |
| Li | Hongkong | 30 m | 7 - 16 | 0.25 | <0.0001 | -164.3<br>(-312.6, -16.1) |
| Gatica | Chile | 30 m | 6 - 14 | 0.46 | <0.0001 | -60.9<br>(-178.2, 56.4) |
| Vandoni | Italy | 20 m | 6 -11 | 0.40 | <0.0001 | -97.8<br>(-223., 28.0) |
| Cacao | Brazil | 30 m | 7 - 12 | 0.46 | <0.0001 | 28.7<br>(-87.1, 144.8) |
| Kahraman | Turkey | 30 m | 6 - 12 | 0.37 | <0.0001 | -52.0<br>(-203.6, 99.5) |
| Jalili | Iran | 30 m | 8 - 16 | 0.29 | <0.0001 | -151.7<br>(-283.1, -20.2) |
| Goemans | Belgium | 25 m | 5 - 12 | 0.30 | <.0001 | -74.2<br>(-206.7, 58.2) |
| Abdulkadir | Nigeria | 30 m | 10 - 18 | 0.10 | 0.749 | 26.7<br>(-113.5, 166.8) |
| Chen* | Taiwan | 30 m | 7 - 17 | 0.96 | <0.0001 | -0.37<br>(-1.03, 0.29) |

*Abbreviations: r=Pearson's correlation coefficient except otherwise indicated; p= p value of linear correlation between observed 6MWD and 6MWD predicted by respective equation; d (lower, upper)= mean bias (mean difference) with lower and upper 95% limits of agreement*
\* Spearman correlation of z scores of observed 6MWD and predicted 6MWD z scores derived with Chen et al's equation
Missing values: for all these analyses n=627, except for comparisons with equations by Jalili for Iranian boys only (n=300), Goemans for Belgian boys only (n=300, boys) and Abdulkadir for Nigerian adolescents only (n=187, 10-11 year-olds).

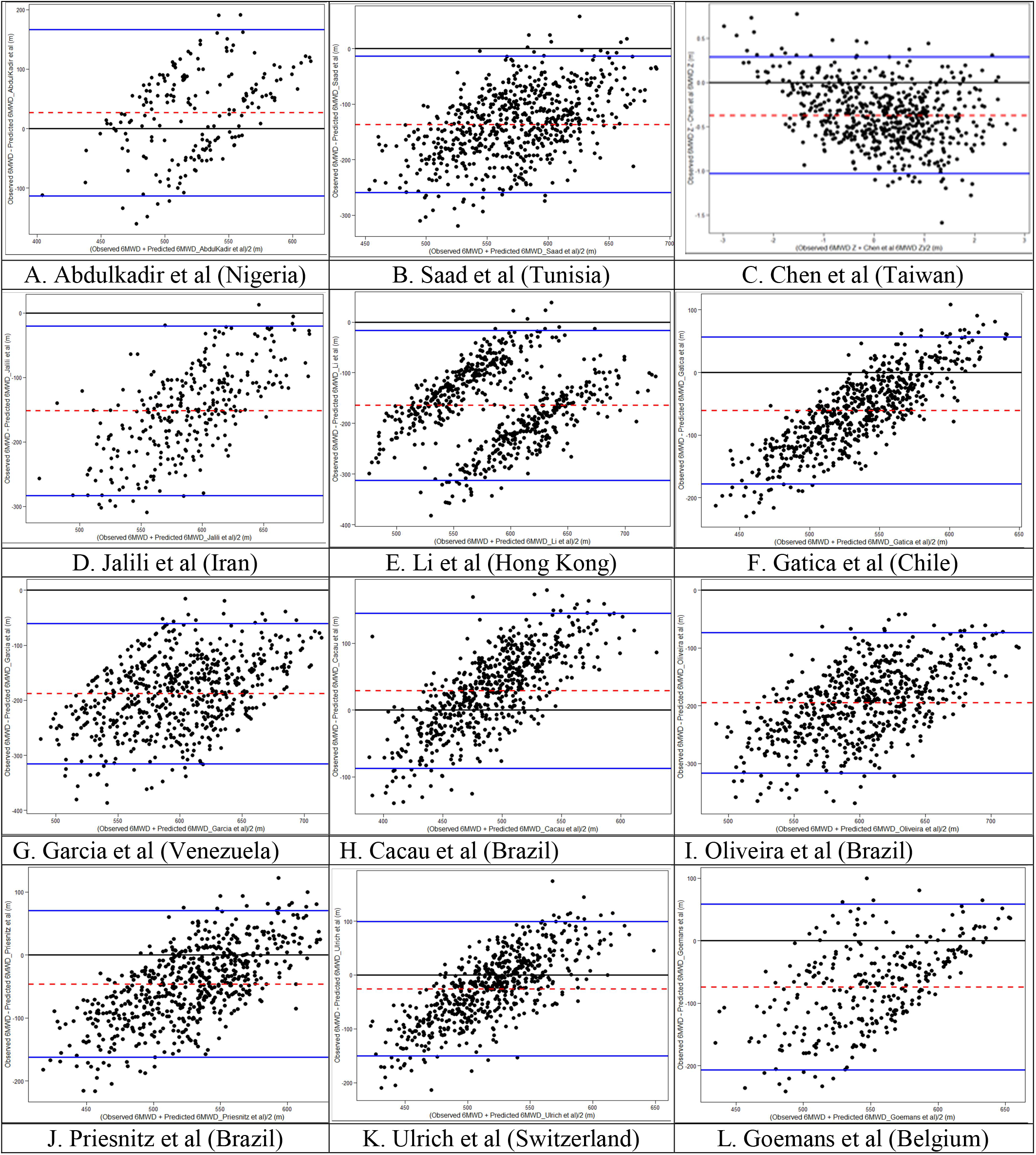
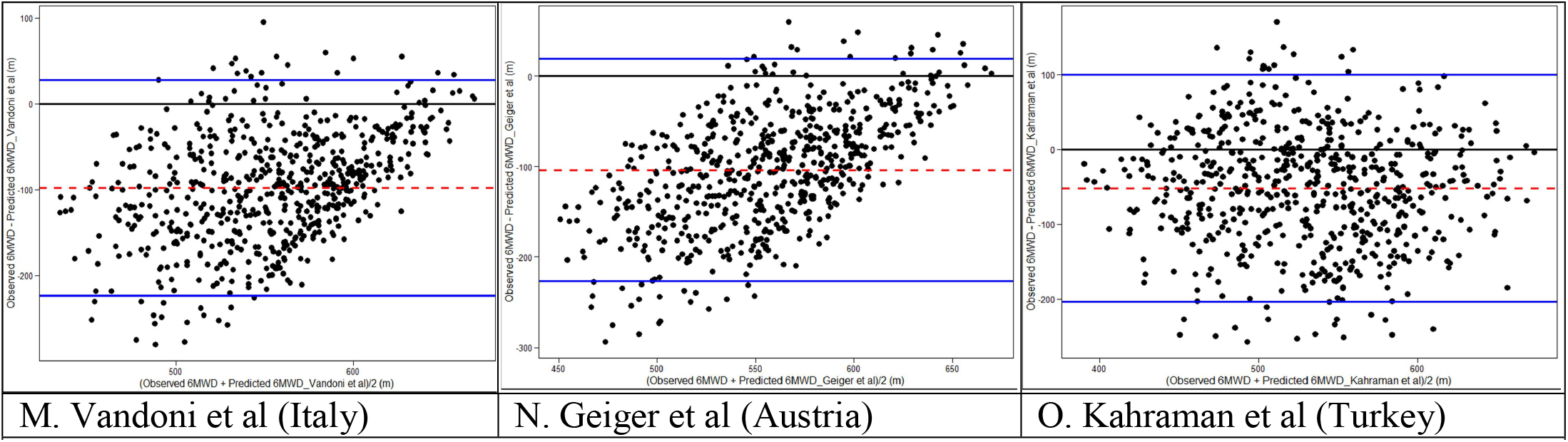
Bland-Altman plots comparing measured 6MWD of Nigerian children aged 6-11 years with predicted 6MWD derived with previously published equations from other populations Keys: dash lines (red) shows the mean difference (bias); outer solid line shows the 95% upper and lower limits of agreement; dots are data points

## DISCUSSION

In this first study of normative standards for the 6MWT of healthy school-age Nigerian children, we report mean 6MWD of 504 m, with age, male sex, change in heart rate (Δ*HR*) and change in systolic blood pressure (ΔS*BP*) independently explaining 25% of the variability; Δ*SBP* as an independent predictor of the 6MWD is a novel finding. Also, none of previously published reference equations satisfactorily predicted our cohort’s 6MWD. Our data adds to existing published ones for Nigerian adolescents(21) and adults,(8, 22) thus completing Nigerian references spanning school-age to adulthood, while also filling an acknowledged paucity of 6MWT data from sub-Saharan Africa.(9, 15)

The 6MWT is a valid and reliable measure of functional exercise capacity widely used in adults and children with disorders like asthma, congenital heart diseases, sickle cell disorders and others. (4, 5, 7, 13). These chronic disorders, and post-recovery phases of some acute illnesses like COVID-19, are associated with impaired functional capacity due to both the underlying pathophysiologic derangements and associated sedentariness.(13, 30) Conceptually, 6MWT measures both *walk capacity* and *functional capacity*-interrelated but distinct concepts in the bio-psychological model of health(2, 3) Thus, 6MWT offer an advantage over CPET which assesses just *walk capacity* which is only a component of an individual’s daily functioning.(3) Also, compared to CPET, 6MWT is easier to perform in sick children because it is self-paced and less exerting.(31)

A mean of 504 m in our study falls within the range of published studies, including systematic reviews by Mylius et al(14) and Cacau et al,(16) of mean 6MWD from healthy children. Compared with previous study by Abdulkadir et al(21) amongst Nigerian adolescents, 10-11-year-old boys and girls in our study walked 544 and 530 m, respectively, while 10-12-year old boys and girls walked 545 m and 491 m, respectively, suggesting similar performances at least in boys. Our 6MWD also compares with 518 m walked by 7-11 year-old North American children.(24)

However, apart from British children aged 4-11 years who walked 470 m,(32) our 6MWD is shorter than published values from North African,(26) European,(23, 33–37) South American(35, 38) and Asian(39) children: the weighted mean 6MWD of European, Asian and South American children were 627m, 604m and 593m, respectively.(15) Summarily, wide differences in mean 6MWD between studies from different, and within countries, can be explained by variations in test procedures, study designs, sample sizes, selection methods, included age groups, and true ethno-racial differences. Methodological modifications are commoner in LMICs mostly due to the use of tracks shorter than the ATS-recommended 30 m.(40) We, and previous authors,(23, 41) conducted the test on 20-meter because shorter corridors are more likely to be available in resource-limited space-constraint settings. Although shorter tracks may result in shorter 6MWD because of frequent slowing down to turn at the ends,(24) this effect seems small especially in children and thus not clinically relevant in tracks at least 15 m long.(13, 24, 41)

Aside the use of shorter track, we adhered to the ATS/ERS recommendations for the 6MWT. For example, we instructed the children to ‘*walk as far as possible*’,(23, 29, 31, 39, 42) rather than ‘*walk as fast as possible*’(11, 24, 32, 34, 35, 38) which could introduce additional motivation to walking faster and longer 6MWD.(4, 43) Also, we tested participants one-at-a-time, rather than in groups or ‘overlapping’(37, 44) which may introduce a competitive motivation to longer distances.(36) We also avoided modifications like the use of measuring wheels,(23) safety chasers(33) or trailing participants to measure *SpO*_*2*_ or *HR*(32) may add additional motication or distract the participant.(32)

Similar to others,(16) we observed a wide range in our participants’ 6MWD, attributable to its self-paced nature variably influenced by individual mood, emotional state or comprehension.(17)(39) However, our large randomly-selected sample size was fairly representative of the target population, evidenced by the narrow confidence intervals.

### Independent predictors

Growth is associated with increasing height, leg length, stride, weight, muscle mass/strength and neuromuscular coordination which all combine to increasing pace and walk distance as a child in age.(18, 33) Thus, in agreement with published pediatric studies,(19, 23, 32, 45) age was the most influential predictor of 6MWD in our cohort.(15) Generally, 6MWD increases with age till about 10-12years, after which a plateau or reduction is observed-attributed to puberty-induced changes in body composition, energy utilization and estrogenic effect.(11, 15, 32, 33, 36, 37, 44, 46, 47) Surprisingly, height was not an independent predictor of 6MWD in our study. Multivariable analysis showed that the observed bivariable correlation between 6MWD and each of height, leg length, *FEV*_*1*_ or PEF was due to the confounding influence of age. Some authors included height, along with age, in their equations but often not stating whether due consideration was given to its potential multi-collinearity with height.

The attainment of longer distance by boys compared to girls in our study, albeit of small effect size, agrees with some paediatric studies,(23, 38, 45, 48) attributable to higher muscle mass, strength, energy levels and activity in boys. (49, 50) In contrast, no significant sex difference was observed in among Tunisian,(26) Thai,(51) Brazilian,(35) Turkish,(19) British and US children.

### Physiological changes during 6MWT

Physiological responses to exercise may reveal cardiopulmonary, hematologic, neuro-musculoskeletal or metabolic abnormalities detectable only when these systems are stressed.(1) Thus, we also provide data on walk-induced physiological changes (SpO2, HR, BP and RPE changes in online supplement) which may serve as comparative standards for exercise-induced changes of sick children before, during and after treatment. For example, 6MWT-associated desaturation, being correlated with nocturnal desaturation,(52) may provide alternative clinically-useful information in children with SCD in the absence of sleep studies facilities. The observed 6MWT-associated changes in HR, SPO2, SBP, DBP and RPE in our study were at most modest and by the 5th minute, all values were close to baseline; similar to reports in British and Brazilian children.(32, 35)

Heart rate change (Δ*HR*), an indicator of the degree of motivation and exertion, is a known independent predictor of 6MWD in both adults and paediatric 6MWT studies.(14, 16–18) The higher the effort exerted during 6MWT, the higher the Δ*HR* and the longer the 6MWD.(18) However, our mean Δ*HR* of 6.5 beats per minute (8% increase from baseline and 45% of predicted maximal heart rate) was small, implying a small degree of exertion similar to 8-10 beats/minute in Turkish(37) children but smaller than others. (33, 39) (35) The higher mean pre-walk, post-walk and 5th minute heart rate values in girls is expected (23, 53) because girls have smaller cardiac size, and hence smaller stroke volume, compared to boys of similar age. They compensate for this smaller heart size with faster heart rate to maintain comparable cardiac output.(49) However, two studies in Indian and Chinese children reported higher mean heart rate values in boys attributed, by the authors, to possible influence of genetics and race on autonomic response to walk.

Inclusion of Δ*SBP* in our model was a unique finding as no published prediction equations in adults or children (after a careful search) have this variable in their equations; 6MWD was not associated with SBP in North African children.(14, 16) Similar to Δ*HR*, Δ*SBP* reflects the effort exerted during an activity.(49) The absence of sex difference in SBP in our study contrasts with an expected exercise-induced physiological response whereby boys have higher walk-induced SBP response due to larger cardiac muscle mass, stronger contractile force and higher stroke volume.(49) The contrasting observation in our study may be due to the low level of exertion as reflected in the small Δ*HR*. The possibility of racial difference in BP response however cannot be excluded. After a careful search, we found no published studies on the physiological response of Nigerian children to walk for comparison.

### Reference equations

Our main equation explained only 25% of our participants’ 6MWD-one of the lowest reported *R*^*2*^-similar to 19% and 25% in Croatian(20) and Italian(54) children, respectively. We paid careful attention to avoid over-fitting our model with variables that violated assumptions of regression such as multi-collinearity to assure goodness-of-fit.(27, 32) Interestingly, the *R*^*2*^ from Nigerian adults was similarly small (30%). Perhaps, anthropometric variables exert less influence on the 6MWD of sub-Saharan Africans compared to other populations, possibly reflecting racial differences in body proportion, composition and movement dynamics. Unfortunately, we found no reference data from other sub-Saharan populations for comparison. An ERS/ATS systematic review in 2014 by Sally et al(7) noted that the *R*^*2*^ in adults ranged from 20-70%.(7) Thus, in both adult and paediatric populations, up to 80% of the variability in 6MWD may remain unexplained by anthro-demographic and physiologic variables due to difficult-to-measure constructs like mood/emotions with wide intra- and inter-individual variations.

### Confounding factors

We cannot explain the reason for the association between 6MWD and school category. Perhaps it indirectly reflects socioeconomic effect mediated through habitual speed of walking rather than anthropometric differences. We did not however include retained school category in the final model as it may be subject to variability from locality to locality. Other anthropometric (BMI, nutritional class), physiologic (SPO2) and psycho-social factors (RPE, physical activity level, ethnicity, time of testing) did not influence the 6MWD of our population. Perhaps, *participation in sports outside school*, rather than objective measurement with accelerometers, may be inadequate as a measure of physical activity. However, this non-association with physical activity agrees with most studies in which PA was measured,(34) except amongst Turkish adolescent where physical activity was an independent predictor of 6MWD.

### Comparison with published reference equations

Similar to previous reports, most published equations significantly over-estimated the performance of our population highlighting need for local reference data.(1, 16) Also, the previous equation from Nigerian adolescents also agreed poorly with our adolescent data despite the narrow bias. This further suggests that 6MWD is also influenced by environmental and geographical factors which may influence habitual speed of walking, aside true ethno-racial, genetic and mood influences.

### Safety

The 6MWT is generally safe, even in ill persons. Although about a third of our cohort reported symptoms such as chest pain while walking, these were clinically non-significant. Among Thai children aged 9-12 years, thigh/leg pain were the commonest complaint (17%) followed by chest discomfort (9%) and dizziness (4%).

### Limitation and recommendations

The absence of repeat testing, as recommended by ATS/ERS updated guidelines for adults, may potentially underestimate our 6MWD. However, practice test exerts comparatively less influence on paediatric 6MWD than adults, thus precluding its necessity.(5, 11, 35) Overall, 7 trained testers performed the 6MWT in our study possibly introducing considerable measurement errors. However, we standardized our procedures and our observed variability are similar to previous reports. Although we did not observe ethnic influence in our data, our findings might not extrapolate to the whole of Nigeria since we conducted it in 1 of over 500 local government areas of Nigeria. Thus, similar study should be conducted in other parts of the country to compare findings and validate our equations.

Unlike Saad et al and Jalili et al, we did not externally validation our equation on a geographically or spatially separate sample as recommended by the TRIPOD consensus statement.(55) However, we used bootstrap procedures for internal validation (our bootstrapped equations demonstrated small biases); this is statistically superior to a split-sample or cross-validation technique reported in some paediatric studies. Because the peak/maximal HR occurred at about the 5th to 6th minute returning rapidly to baseline within one minute of stopping to walk(32, 41), especially in children,(41) our use of intermittent, rather than continuous,(4, 5) measurement of heart rate may have underestimated the true peak/maximal and immediate post-walk HR values and consequently the predictive contribution of Δ*HR* in our equation. Hence other researchers should employ continuous heart rate monitoring during 6MWT. To further reduce variability in 6MWT procedural methodologies and its influence on the variability in reported 6MWD, there is need for paediatric-specific guidelines for 6MWT.

## CONCLUSIONS

Nigerian school-aged children achieved 6MWD shorter than most published reference values, except those of British children. The independent predictors of the walk distance were age, male sex, ΔHR and ΔSBP – the latter a unique finding; all explaining only 25% of the 6MWD variability. Published equations from North Africa and non-African countries mostly over-estimated the 6MWD but there were none from other sub-Saharan populations to compare with. Till date, large variability in reported 6MWD and its predictors precludes the development of a global reference equations for the 6MWT as was possible with the Global Lung function initiative.(14–16, 56)

## Supporting information

CoI form

TRIPOD Checklist

## Data Availability

All data produced in the present work are contained in the manuscript and supplementary file, but further details are available on request from the lead author

## ACKNOWLEDGEMENT

The authors acknowledge Dr ‘Titi Ogunlana who deputised for the PI especially while away from administrative clinical responsibilities; the Research Assistants (namely Raphael Adebayo, BSc ‘Niyi Olusanya, Ebere David, ‘Tobi Yekini and Chibuzor Nwosu, Bsc) for being part of this project; Chris Ubuane for helping to procure some of the devices and accessories from the UK. Our thanks also goes to the Lagos State Universal Basic Education Board (SUBEB) and the Lagos State Ministry of Education for granting permission to conduct the study, Head Teachers, Proprietors and School Staff, the amiable and enthusiastic pupils, along with their parents/guardians-who participated in the study are specially appreciated.

## Footnotes

part of this data has been submitted as a dissertation to the Faculty of Pediatrics of the West African College of Physicians and as conference abstract

## AUTHORS’ ROLES

POU: conceptualization, design, data acquisition, statistical analysis, initial draft, revision, final draft

OAA: design, training, revision, final draft

OAA: data acquisition, revision, final draft

GA: data acquisition, revision, final draft

CIA: data acquisition, revision, final draft

MOK: data acquisition, revision, final draft

OAA: data acquisition, revision, final draft

MOA: data acquisition, revision, final draft

BAA: conceptualization, design, initial draft, revision, final draft, supervision/senior author

FON: conceptualization, design, statistical analysis, initial draft, revision, final draft, supervision/senior author

