## Supplementary File for "Development of reference equations for the six-minute walk distance of school-aged Nigerian children"

TITLE:

### METHODS

#### STUDY AREA

We conducted the study in Ikeja Local Government Area (LGA) of Lagos State, South-West Nigeria. Ikeja LGA is one of the 20 LGAs in Lagos State; it is the capital of the state and reflects the cosmopolitan characteristics of the state and of Nigeria, with its inhabitants comprising various ethnic groups, predominantly the Yorubas and Igbos. Ikeja LGA had three educational districts during the study period.

#### STUDY DESIGN

The study was observational and cross-sectional.

**STUDY POPULATION**

This consisted of primary school pupils aged 6-11 years from the three educational districts of Ikeja Local Government Area. Children younger than six years of age were not included because they may be unable to follow the instructions required for the test.

**Inclusion criteria**

1. Apparently healthy Nigerian primary school pupils aged six to eleven years;

2. Satisfactory completion of a screening questionnaire by parents/guardian;

3· Cognitive ability to understand and obey procedural instructions;

**Exclusion criteria**

1. Refusal of consent by parents and/or assent by pupil;

2. Diagnosis or history or physical finding suggestive of any acute or chronic disorder like sickle cell disease, anemia, cardiopulmonary diseases, asthma, neurologic or musculoskeletal disorders;

3. Hospitalization in the last three months;(1)

4. History or sign of any acute illness, including respiratory tract infections, within the last three weeks;(2)

5. Current use of any medications that could potentially affect walk test (e.g. anticonvulsants) or drug therapy for acute illnesses (e.g. antimalarial);

6. Elevated blood pressure ($\geq 90^{th}$ percentile for age, sex and height);(3, 4)

7. Resting oxygen saturation of at least 95%;(5)

7. Abnormal spirometry ($FEV_1$ values < 80% of predicted for sex, age and height)(6)

### SAMPLE SIZE

The minimum sample size was determined using the standard formula for estimation of single mean:(7) $N = (z^2 \text{ x } \alpha^2)/t^2$ where N = sample size, $z$=standard deviation at 95% confidence interval (1.96); $\alpha^2$=population variance and $t$ = level of precision. From a previous study of 200 African (Tunisian) school pupils, the mean (± standard deviation) for the 6MWD was 700 ± 73).(8) Assuming a 1% degree of precision of the mean and confidence coefficient of 95%,

$$N = [(1.96)^2 \text{ x } (73)^2]/(7)^2 = 417$$

Thus we needed at least 417 pupils to estimate the mean 6MWD of Nigerian children within 1% precision (objective 1). However, we exceeded this minimum size in order to enhance the precision of our statistical estimates and prediction equations (other objectives).(9)

### SAMPLING TECHNIQUE

We recruited participants from selected primary schools with a multi-stage sampling technique thus:

*Stage 1: Sampling frame*

Our sampling frame consisted of a list of the registered 32 public and 176 private schools within the Ikeja LGA of Lagos State obtained from the Lagos State Universal Basic Education and Ministry of Education, respectively.

*Stage 2*: *Selection of schools by simple random sampling*

The names of all the schools were written on separate pieces of paper (separately for each of public and private schools). We asked an observer who was not part of the research team randomly picked five public schools and ten private schools which were written on pieces of papers (we arbitrarily selected this initial number of schools in the absence of data on the total number of enrolled pupils in each of the schools to guide the number of schools from which the sample would be obtained). The selected schools were visited consecutively. Schools that declined participation or that had no appropriate testing sites (hard-surfaced, straight course in a corridor, hallway or walkway of at least 20m long with allowance for additional spaces for turning at the two ends) were replaced with the next consecutive school on the main lists.

*Stage 3*: *Selection of classes and participants by random sampling*

In each selected school, participants were recruited from each of six classes (primary one to six). Where there was more than one arm per class, one of the two or more arms was randomly selected by balloting. In each selected class, with all pupils seated, every *n*th pupil was selected where '*n*' is the number of pupils in the class divided by the number of students to be selected from the class. For the public schools a minimum of 17 pupils (9 girls, 8 boys) were required in each class and for private schools, 22 pupils (11 girls, 11 boys). Where the number of recruited pupils in a school was inadequate, more pupils were recruited from the next selected school. Recruitment thus continued consecutively from school to school.

**ETHICS**

The study was approved by the Health Research & Ethics Committee of the Lagos State University Teaching Hospital (LREC/10/06/486) and was officially permitted by the Lagos State Universal Basic Education and Ministry of Education. We also obtained written parental consent as well as each pupil's assent before enrolment and participation.

**Action on excluded subjects**

All the children with symptoms, signs and abnormal screening tests findings had written reports sent to their parents/guardians through the school authorities. The written reports were supplemented with e-mails, phone calls, short message tests or 'whatsapp messages' sent directly to the parents/guardians. They were invited to bring their wards for further evaluation at the paediatric Out-patient clinics of LASUTH or any other hospital of their choice (some of the pupils were subsequently evaluated for asthma, adenoidal hypertrophy, missed beat and elevated blood pressure).

#### TRAINING OF INVESTIGATOR AND RESEARCH ASSISTANTS

Before commencement of field work, the principal investigator and a research assistant were trained and standardized on the conduct of the 6MWT by the second author- a cardiopulmonary physiotherapist who had prior wide experience in the clinical and research administration of the 6MWT.(10, 11) The PI subsequently trained other research assistants on the 6MWT as well as on anthropometric measurements, physiologic measurements (blood pressure measurement, pulse oximetry, perceived exertion measurements), data collection and recording. Four research assistants (two with university degrees and two with secondary school certificates) assisted the investigator with data collection over a time period ranging from four to twelve months, along with six doctors (five resident doctors and one consultant pediatrician).

#### Pre-testing/pilot

After the training, the PI pretested the 6MWT on a patient in the hospital and then on six pupils of the first school visited to gather further experience with the test administration and modify the sequence of data collection; their data were not included in this analysis.

#### Study duration

Field work was conducted from April 2016 to June 2017 during school sessions, excluding examination periods. The number of pupils recruited per day ranged from 2 to 20 pupils and varied from school to school. Tests were conducted during school hours (8.00 PM to 2.30 PM).

### PRE-TEST EVALUATION

During the first visit to each school (after approval), the investigating team was introduced to the pupils class-by-class. The PI explained the nature and purpose of the study explaining its purpose, methods, and safety. Questions on the test were entertained from teachers and pupils and answered. We then gave each selected assenting pupil a written statement of consent, with a screening questionnaire attached, to be delivered to their parents/guardians. The questionnaire consisted of questions on the child's demographic characteristics (age, sex, school name, class, address, parents/guardians phone/email), health-screening questionnaire (physical activity readiness questionnaire-child, PARQ-child)(12, 13) and other questions to exclude features of sickle cell disorder. Consenting parents were required to sign the completed questionnaire and return them through their wards.

At the second visit, we retrieved the consent forms and questionnaires from the pupils and reviewed them for parental permission, completeness and eligibility. After further explaining the purpose of the study in simple terms to each pupil whose parents consented and satisfactorily completed the questionnaire with no exclusion criterion, we obtained written assent from each pupil in addition to parental consent. Verbal assent was however obtained from all the participants. Pupils who declined assent, despite parental consent, were thanked and asked to return to class. We discouraged class teachers from trying to coerce pupils to participate, stressing absolute voluntariness.

#### Physical evaluation

We sought for history of frequent exercise-induced chest pain and/or dizziness, palpitation at rest and current lower limb pains or injuries from assenting pupil to ensure there were no symptoms suggestive of any acute or chronic condition that could impair test performance.(3) The PI, or any of the 5 assisting doctors, then conducted a *physical examination* on each pupil, consisting of checking for pallor (conjunctival, buccal and palmar), jaundice, cyanosis, finger-clubbing, pedal edema, gait abnormalities, as well as abnormal cardiac or breathe sounds.

**Thermometry**

We excluded present fever (temperature above 37.5˚C.(14)) in each pupil with a calibrated digital non-contact forehead infrared thermometer (FDK®, Fudakang Industrial Co. LTD, China).

**Screening spirometry**

We used a hand-held paediatric digital peak flow meter (asma-1™; Vitalograph, England) that satisfies ATS accuracy standards to measure each pupil's peak expiratory flow rate (PEF) and forced expiratory volume in one second ($FEV_1$).(15, 16) A disposable one-way mouthpiece with filter (*SafeTway*® mouthpiece; Vitalograph, England) was attached to the device and used for individual participant to avoid cross-contamination.(15) Spirometry was done according to American Thoracic Society/European Respiratory Society (ATS/ERS) guidelines.(17) With the participant in standing position and head slightly elevated instruction was given to inhale rapidly and fully. At the same time, the participant placed the mouth-piece in the mouth with the lips around it, exhaled rapidly and forcefully until no more air could be expelled. These steps were repeated at least two more times; pupils with difficulty in following the steps had further demonstration and attempt at the procedure for a maximum of eight times in total. We recorded the PEFR and FEV1 values displayed at the end of each attempt. We then derived their percentage predicted values using local prediction equations by Faleti.(6) FEV1 values less than 80% of predicted for sex, age and height was regarded as abnormal and such pupils were excluded and invited to LASUTH for further clinical evaluation.

**Anthropometry**

*Height*

We measured height to the nearest 0.1cm with a stadiometer (Prestige® Height measuring stand, Haerdik Medi Tech, India) without shoes and with the heels and back against the height meter while looking straight ahead with the lower borders of the eye sockets in the same horizontal plane as the external auditory meatus (Frankfort position).(18) The measurer then moved the slider down to touch the participant's head and measured the height at the same horizontal level.

*Weight*

We measured weight was measured to the nearest 0.1kg with an electronic weight scale (Omron® HN283, OMRON Healthcare Co LTD, Kyoto, Japan) with sensitivity of 100g. The pupils were weighed in their school uniforms without shoes, socks, cardigan or sweater and with all pockets emptied out.(18)

*Lower Limb length* (*leg length)*

Leg length was measured on the right lower limb with an inelastic flexible tape to the nearest 0.1cm with the participant in standing position from the anterior superior iliac spine (ASIS) to just below the medial malleolus.(19, 20)

*Chest circumference (CC)*

Chest circumference was measured with an inelastic tape measure (sensitivity of 0.1cm) to the nearest 0.5cm at the xiphisternal junction at the end of tidal inspiration (without exposure other than removal of extra wears such as cardigans, ties)(21).

The mean of two readings of all the above-mentioned anthropometric measurements were obtained.

**Blood pressure (BP)**

We measured BP using an automatic digital blood pressure monitor (Omron® 705IT, OMRON, Netherlands) validated for clinical use in children and adolescents.(22) The blood pressure reading was measured during a 5-10 minutes rest before the walk test. With the right arm at the heart level after at least five minutes of rest, a cuff appropriate to the participant's arm circumference was used; the length of the inflatable bladder covered at least 80% of the mid arm circumference and the width about 40-50% of the mid arm circumference.(4) The mid-arm circumference was measured midway between the acromion of the scapula and the olecranon of the ulnar with the elbow held at 90˚ angle.(18) The cuff was applied 1-2cm above the elbow according to manufacturer's manual and then inflated by pressing the start button. The cuff inflates automatically and gives systolic and diastolic pressures which were recorded in mm of mercury (mmHg). We used a smart phone application (Pediatric Assistant®)(23) to derive on-the-spot percentile scores for each BP reading based on the 2004 Centre for Disease & Prevention (CDC)'s values for the participant's age, sex and height.(4) Readings persistently at or above the $90^{th}$ percentile were regarded as abnormal and such pupils were excluded from further participation and invited to LASUTH for further clinical evaluation.

**Oxygen saturation (SpO2) and heart rate (HR)**

Oxygen saturation ($SpO_2$) and heart rate (HR) were measured on the left thumb with a finger pulse oximeter (Nonin® Onyx Vantage 9590, Nonin Medical Inc., Minneapolis, MN, USA). The $SpO_2$ and HR values were recorded and expressed in percentages (%) and beats per minute (bpm), respectively.

**Rating of perceived exertion (RPE)**

Each participant was shown a pictorial RPE scale by Cassady et al(24) and then asked to point to the picture that best describes *how tired he or she felt* (perceived level of exertion) before the 6MWT. The corresponding numerical value was then recorded as the prewalk or baseline RPE.(24)

### SIX-MINUTE WALK TEST

#### Test site

In each selected school, we identified and selected an outdoor flat, smooth-surfaced or paved corridor, hallway or walkway with minimal traffic as the test site, sometimes condoned off with red or white tapes to minimize intrusion by playing children during break periods. A 20-meter straight course was marked out on the surface of the floor with a red-colored tape strapped to the floor. The tape was marked at every one meter further sub-divided and marked at every 5 cm to ensure ease of measurement. The starting (0 m mark) and endpoints (20 m mark) were clearly demarcated as turn-around points with small-sized brightly-colored traffic cylinders.(3, 25, 26)

#### Test procedure

The 6MWT was administered according to ATS/ERS(3, 27) guidelines by two persons at a time: a *test instructor* (the investigator or any of the research assistants or assisting doctors trained for 6MWT administration) who evaluated the participant's RPE, gave pre-walk instructions, ensured safety of footwear, administered the 6MWT including minute-by-minute encouragements and timed the test; and a *test assistant* (any of the other research assistants) who counted the laps with the aid of a lap counter sheet and also measured the immediate and $5^{th}$-minute post-walk BP, $SpO_2$ and HR.(3)

After about 5-10 minutes of rest on a seat close to the starting line (anthropometric, blood pressure, pulse oximetry and spirometry measurements were done during this period), the instructor asked the participant to stand at the starting line of the track. ATS standardised test instructions were then read to the participant, stating that the test aimed to determine *how far* *s/he can walk in six minutes* without running, jumping, hopping, or skipping, but s/he could rest or stop during the test if but the timer would not be stopped. The instructor gave opportunity for question or clarification and ascertained the participant's comprehension before the walk. All instructions before and during the tests were communicated to the pupils in simple English. For pupils with poor understanding of English, the local dialect (Yoruba) was used. Where effective communication of instruction was still difficult, the participant was excluded. One participant was tested at a time to avoid competition.

Thereafter, the instructor demonstrated the test by walking *briskly* to and fro the end of the track.(3) After this, another opportunity was given to the participant to ask question. The instructor then took the RPE reading and positioned the participant beside the 0m mark of the tape and asked to START walking. At the same time a stopwatch is started by the instructor. Only ATS Standardised phrases of encouragement, with an announcement of the time remaining were read out to the participant every minute. During walk test, the instructor stay somewhat close to the walking participant without pacing, trailing or obstructing the participant, while announcing the encouragements and time in "even tone of voice".(3)

| Table E1: ATS STANDARDIZED ENCOURAGEMENT FOR THE 6MWT |
|---|
| After the first minute: "***You are doing well. You have 5 minutes to go***." <br> When the timer shows 4 minutes remaining: "***Keep up the good work. You have 4 minutes to go***." <br> When the timer shows 3 minutes remaining: "***You are doing well. You are halfway done***." <br> When the timer shows 2 minutes remaining: "***Keep up the good work. You have only 2 minutes left***." <br> When the timer shows only 1 minute remaining, "***You are doing well. You have only 1 minute to go***." |

Reference(3)

The *test assistant,* sitting on a second chair close to the starting line, counted the number of laps walked by ticking a box on the *lap counter* worksheet on the participant's data collection form for every completed lap. One complete lap consisted of walking from the starting line to the end of the 20m long track, turning around the 20 m mark demarcated by a traffic cylinder and walking back to the starting line. Thus, a lap equaled 40m.(28)

At the end of the sixth minute, the participant is told to STOP walking and remain at the point reached. The instructor quickly walked to the participant, administered and recorded the RPE. At the same time, the test assistant offered the participant a seat at the point reached, measured and recorded the post-exercise BP, $SpO_2$ and HR as immediately as possible within one minute of stopping the walk. The test assistant also measured the 6MWD by noting the point along the tape where the participant's most forward foot stopped; the distance from the starting point to this point is recorded as the *additional uncompleted lap*.(1) The 6MWD equals *40 X (number of laps) + the additional uncompleted lap*.

**5th-minute post-Test evaluation**

After taking the post-exercise readings, the participant was asked to rest on a seat close to the starting point for five minutes.(3) While resting, we asked for specific symptoms felt while walking: chest pain, chest tightness, dizziness, palpitation, leg pain and cramps. Where there was presence of any symptom, such symptom was further characterized by asking for severity, previous occurrences during play or sport activities and associated symptoms. At the end of the five-minute post-walk rest, we repeated the HR, $SpO_2$ and BP. Sometimes, we aborted the $5^{th}$-minute repeat measurements when pupils needed to resume class lessons immediately.

### QUALITY CONTROL MEASURES: (CALIBRATION AND STANDARDISATION)

The two electronic sphygmomanometers were periodically measured against aneroid sphygmomanometer used in the Paediatric Nephrology Clinic of LASUTH. The digital sphygmomanometers maintained a reading of about ±5mmHg with the mercury sphygmomanometer.

The investigator also regularly cross-checked anthropometric measurements taken by the assistants to ensure acceptable reliability (Inter-rater reliability for chest circumference, lower limb length and height were respectively 0.997, 0.826 and 0.992).

Heart rate of a non-participant was periodically counted manually over one minute and compared with the heart rate obtained with the pulse oximeter to ensure accuracy. The pulse oximeters HR reading was within ± 6 beats per minute of manually counted HR on most days. It also maintained close reading to HR reading of the digital sphygmomanometer.

The digital weight scales were also periodically compared with a beam balance in the Paediatric Emergency of the Hospital (LASUTH) to ensure proper functioning and accuracy.

The spirometer, according to the manufacturer, required no calibration.(15) However, the investigator periodically used it to check personal $FEV_1$ and PEF: it maintained a consistent reading.

#### PARTICIPANTS' SAFETY(3, 26)

1. Participation was entirely voluntary without coercion by the research team or school staff;
2. Class teachers, including those supervising pupils' physical education or activities, decided when to release pupils and controlled their orderly release from classes (once lessons resumed, pupils were sent back to class till further free periods);

3. Pupils with exclusion criteria were politely excluded and invited to LASUTH for further clinical evaluation through a written report sent to parents/guardians;
4. Tests were conducted within school premises, preferably close to the administrative block and within view of school staff;
5. Safe test sites were carefully selected in each school by avoiding corridors with bumps or craters that could trip walking pupils. Schools without smooth-surfaced test sites were skipped and replaced with others;
6. Basic resuscitation devices were made available on-site (manual bag and mask, battery-operated nebulizer, salbutamol nebules and oxygen source);(3)
7. Pupils were not allowed to carry out the test in slippers, high-heel shoes or barefoot(25);
8. Participants were free to slow down, rest or stop during the test in the event of any intolerable discomfort;
9. Disposable, single-user mouth-pieces with in-built one-way filter were attached to the spirometer for the screening spirometry. The spirometer device was cleaned periodically with methylated spirit according to manufacturer's instruction;(15)
10. Tape measures were wiped with methylated spirit to limit cross-infection;(18)
11. Digital electronic, rather than mercury-containing, sphygmomanometer and thermometer were used to avoid risk of mercury spillage and toxicity;
12. We sent written report of abnormal findings, with advice on appropriate referral steps, to parents/guardians.

**DATA MANAGEMENT**

We recorded participants' data obtained with a self-designed data collection form. WHO's AnthroPlus® (version 1.0.4, WHO, Geneva 2009) was used to derive each participant's age in years (to one decimal place) from the date of birth and anthropometric z-scores [BMI-for-age z-score (BAZ), height-for-age z-scores (HAZ) and weight-for-age z-score (WAZ)]. WAZ was derived only for children up to 10.0 years as WHO Nutritional classification does not use WAZ scores for children older than 10.0 years; rather BAZ and HAZ are used for nutritional classification of children older than 10.0 years of age.(29) Classification of participants' nutritional status was based on WHO criteria: BAZ ≥ -2 to +1 was defined as normal weight, BAZ > +1 to ≤ +2 as overweight, BAZ > +2 to +3 as obese, BAZ > +3 as severely obese, and BAZ < -2 as thin. WAZ <-2 was defined as underweight and HAZ < -2 as stunted.(29)

We extracted data from the data collection forms into Microsoft® Excel 2013 Spreadsheet (Microsoft Inc., USA) which was then imported, for statistical analyses, into Bluesky Statistics® version 7.30 (BlueSky Statistics LLC, Chicago, IL, USA; https://www.blueskystatistics.com) and JASP Statistics® version 0.14 (University of Amsterdam, the Netherlands; https://jasp-stats.org)- both are free open-source graphical user interphases (GUI) for R.

**Univariable analyses**

During data cleaning, we checked for outliers or implausible values; these were corrected when due to data entry errors. Persistent outliers were included in analysis and their influence on subsequent models was assessed with Cook's distances. After data cleaning, we computed the following variables:

1. Body-mass index (BMI)= Weight (Kg)/ Height (m)$^2$
2. Six-minute walk distance = (40 x Number of completed laps) + additional uncompleted lap
3. Heart rate change ($\Delta HR$) = difference between post-walk heart rate (*PostHR*) and pre-walk heart rate (*PreHR*)
4. Predicted maximal HR ($PredHR_{max}$) =208 – (0.7 x age in years)(30)

5. Percentage of predicted maximal heart rate (*%*$PredHR_{max}$) = ($HR_{max}$/ $PredHR_{max}$)*x100%* [where the post-walk heart rate was defined as equivalent to the maximal heart rate ($HR_{max}$)]
6. $SpO_2$ change ($\Delta SpO_2$) = difference between post-walk $SpO_2$ ($PostSpO_2$) and pre-walk $SpO_2$ ($PreSpO_2$)
7. SBP change (Δ*SBP*) = difference between post-walk SBP (*PostSBP*) and pre-walk SBP (*PreSBP*)
8. DBP change (Δ*DBP*) = difference between post-walk DBP (*PostDBP*) and pre-walk DBP (*PreDBP*).
9. RPE change (Δ*RPE*) = difference between post-exercise RPE (*PostRPE*) and pre-exercise RPE (*PreRPE*)

Shapiro-Wilk test and visualization of scatter and Q-Q plots were used to assess the distribution of continuous variables.(9) Categorical variables such as sex, age groups, school category (public versus private), ethnic group and sport activity were expressed as frequencies (percentages, %) while continuous variables such as 6MWD were expressed as means with standard deviation (SD) and 95% confidence intervals (95%CI) for normally-distributed variables, or median with interquartile range (IQR) when significantly skewed.

**Bivariable analyses**

We used Student t-test (or Mann-Whitney u-test if skewed) to compare continuous variables between two independent categories such as sexes (girls vs boys), school categories (public vs private), time of testing (AM vs PM) and participation in sport activity outside school hours (yes vs no). One-way Analysis of Variance (ANOVA), with linear trend test was used to assess the association between the 6MWD and age group.(9) We used 1-way and 2-way repeated-measure ANOVA to assess changes in each of *HR*, $SpO_2$, *SBP* and *DBP* over time (at baseline, immediate and 5th minute) in the total sample and between the sexes, respectively. For the RPE, we used Wilcoxon-signed rank test to assess change in RPE from baseline to post-walk. Some pupils could not conclude the 5th-minute measurements because they had to return to classes. Missing data points for the 5th-minute values of *HR*, $SpO_2$, *SBP* and *DBP* were handled with *single imputation function* in Bluesky statistics®).(31) However, the inputted variables were only used for the repeated-measure ANOVA analyses. During 2-way ANOVA, we checked for violations of assumptions of normality of variances, sphericity (violation was corrected by adopting the Huynh-Feldt correction), equality of variances and interactions. Multiple comparisons during 2-way ANOVA and linear correlation were corrected with Bonferroni-Holms' method.

The effect sizes of statistically significant comparisons were assessed with Cohen's d for t-tests (trivial: < 0.1, small/weak: 0.2, medium/moderate: 0.5 and large/strong: 0.8) and ANOVA (trivial: < 0.1, small/weak: 0.1, medium/moderate: 0.3 and large/strong: 0.5); rank biserial correlation for Mann-Whitney U-test and Wilcoxin signed-rank test (trivial: < 0.1, small: 0.1, medium/moderate: 0.3 and large/strong: 0.5); partial eta squared ($\eta^2p$) for ANOVA and repeated-measure ANOVA (trivial: < 0.1, small: 0.01, medium/moderate: 0.06 and large/strong: 0.14)(32) Pearson's correlation coefficient, or Spearman's rho if skewed, was used to assess the linear relationship between the 6MWD and each continuous variable (trivial: < 0.10, small: 0.10, medium/moderate: 0.30 and large/strong: 0.50).(32) The linearity of the association between the 6-MWD and each of these variables was also assessed with scatterplots.

#### MULTIVARIABLE ANALYSIS

We used stepwise multivariable linear regression models to derive equations with the *least number of easy-to-measure variables, highest regression coefficient, highest coefficient of determination ($R^2$) and the least standard error of estimate (SEE)*. Variables with significant association with the 6MWD in preceding bivariable analyses were first *stepped* into an initial exploratory model using default settings in JASP (significance levels of 0.15 and 0.05). The initial exploratory model was assessed for goodness-of-fit and non-violation of statistical assumptions of multivariable linear regression, and further simplified in subsequent stepwise procedures. During model development, we assessed goodness-of-fit with R square *($R^2$),* adjusted $R^2$, multiple correlation coefficient ($R$), standard error of the estimate (SEE), standard errors of the unstandardised beta coefficients and p-values.(9) We ensured: absence of multicollinearity (defined as variable inflation factor $\geq 4$);(9) absence of heteroscedasticity (by visually inspecting plots of standardised residuals against standardised predicted values for uniform distribution of data points along changing predicted values);(33) normality of residuals (bell-shaped histogram of the standardised residuals); normal probability plot (residuals dots lying approximately on the line in the graph); and absence of influential outliers or extreme cases (Maximum *Cook's distances* less than 1.00).(9, 33)

**Internal validation**

We used bootstrapping techniques with 5,000 iterations (JASP Statistics) for internal validation of the equations using the bias. We present the bootstrapped equations.

**Comparison with previously published equations**

Predicted 6MWD derived with previously published equations were compared with our sample's measured/observed 6MWD using linear correlation and Bland-Altman technique for measurement of agreement between two methods of measurement.(34) Bland-Altman technique included the following steps:

- The actual distances walked by each participant (observed 6MWD) was regarded as the 'gold-standard' measure while the 6MWD predicted by the published equation were regarded as the second measure (predicted 6MWD);

- the difference between the observed 6MWD and predicted 6MWD (*6MWD difference*) was calculated for each participant;
- The mean of the 6MWD differences (mean difference or bias, *d*) and its standard deviation (*s)* were calculated;
- The 95% upper and lower limits of agreement (LoA) were, respectively, calculated as *d* + 1.96*s* and *d-1.96s*;
- The 6MWD difference of each participant was plotted on the Y-axis while the mean 6MWD was plotted on the X-axis of a scatter plots;
- The mean difference (*d*), the upper LoA and lower LoA were also plotted;
- Interpretation of a Bland-Altman plot requires that a clinically acceptable limit of agreement (upper and lower limits) be determined *a priori*.(34) With respect to the 6MWD, a universally acceptable LoA may be impracticable as it may vary widely from disease to disease. However, we considered that LoA that do not exceed the previously reported minimally detectable change of 50 m(1) among healthy children was narrow enough for acceptable agreement.
- We also inspected the plot visually for proportionate bias

### RESULTS

Table E2. Comparison of anthropometric variables between girls and boys

Height (cm)

| Age | Girls | | Boys | | All | *p* |
|---|---|---|---|---|---|---|
| | **N** | **Mean (SD)** | **N** | **Mean (SD)** | **Mean (SD)** | |
| 6 | 49 | 117.2 (5.8) | 38 | 116.3 (5.6) | 116.8 (5.7) | 0.479 |
| 7 | 54 | 122.5 (6.6) | 38 | 120.8 (5.0) | 121.8 (6.0) | 0.184 |
| 8 | 70 | 126.6 (7.0) | 70 | 127.9 (5.9) | 127.2 (6.4) | 0.265 |
| 9 | 59 | 132.7 (10.1) | 62 | 133.6 (7.9) | 133.1 (9.0) | 0.576 |
| 10 | 62 | 136.9 (8.0) | 58 | 138.1 (8.4) | 137.4 (8.2) | 0.426 |
| 11 | 33 | 142.4 (8.8) | 34 | 136.7 (8.6) | 139.5 (9.1) | 0.101 |

Weight (kg)

| Age | Girls | | Boys | | All | *p* |
|---|---|---|---|---|---|---|
| | **N** | **Median (IQR)** | **N** | **Median (IQR)** | **Median (IQR)** | |
| 6 | 49 | 21.6 (19.7, 24.4) | 38 | 21.2 (19.7, 23.1) | 21.4 (19.6, 23.5) | 0.653 |
| 7 | 54 | 24.6 (22.0, 29.7) | 38 | 23.3 (21.5, 27.4) | 23.6 (21.8, 28.2) | 0.288 |
| 8 | 70 | 26.9 (23.7, 31.9) | 70 | 28.8 (25.2, 32.6) | 27.5 (24.3, 32.4) | 0.097 |
| 9 | 59 | 31.1 (25.7, 38.5) | 62 | 30.1 (27.0, 34.4) | 30.3 (26.3, 36.8) | 0.608 |
| 10 | 62 | 32.0 (28.5, 37.1) | 58 | 33.2 (28.3, 40.5) | 32.7 (28.5, 38.9) | 0.596 |
| 11 | 33 | 37.6 (32.8, 45.4) | 34 | 33.7 (28.7, 38.1) | 35.5 (29.6, 40.8) | 0.052 |

BMI ($kg/m^2$)

| Age | Girls | | Boys | | All | *p* |
|---|---|---|---|---|---|---|
| | **N** | **Median (IQR)** | **N** | **Median (IQR)** | **Median (IQR)** | |
| 6 | 49 | 15.5 (15.0, 17.4) | | 16.0 (15.2, 16.5) | 15.8 (15.1, 16.7) | 0.864 |

| | | | | | | |
|---|---|---|---|---|---|---|
| 7 | 54 | 16.1 (15.4, 18.3) | 38 | 16.1 (15.2, 18.0) | 16.1 (15.4, 18.3) | 0.643 |
| 8 | 70 | 16.8 (15.7, 18.7) | 70 | 17.1 (16.0, 19.5) | 16.9 (15.9, 19.1) | 0.231 |
| 9 | 59 | 17.2 (16.1, 20.0) | 62 | 16.9 (16.0, 18.1) | 17.0 (16.0, 19.0) | 0.416 |
| 10 | 62 | 17.6 (16.4, 18.8) | 58 | 17.4 (15.9, 20.0) | 17.5 (16.3, 19.5) | 0.916 |
| 11 | 33 | 18.1 (17.2, 20.0) | 34 | 18.2 (16.9, 19.7) | 18.1 (17.1, 19.9) | 0.590 |

Lower limb length (cm)

| **Age** | Girls | | Boys | | All | ***p*** |
|---|---|---|---|---|---|---|
| | **N** | **Mean (SD)** | **N** | **Mean (SD)** | **Mean (SD)** | |
| 6 | 49 | 71.1 (4.8) | 38 | 69.8 (4.4) | 70.5 (4.6) | 0.22 |
| 7 | 54 | 75.3 (5.4) | 38 | 72.4 (4.4) | 74.1 (5.2) | 0.007 |
| 8 | 70 | 77.1 (5.1) | 70 | 77.5 (4.4) | 77.3 (4.8) | 0.611 |
| 9 | 59 | 81.7 (7.9) | 62 | 81.8 (5.6) | 81.8 (6.8) | 0.958 |
| 10 | 62 | 84.8 (6.5) | 58 | 84.8 (6.6) | 84.8 (6.5) | 0.991 |
| 11 | 33 | 88.2 (6.7) | 34 | 82.8 (8.2) | 85.4 (7.9) | 0.004 |

Chest circumference (cm)

| **Age** | Girls | | Boys | | All | ***p*** |
|---|---|---|---|---|---|---|
| | **N** | **Mean (SD)** | **N** | **Mean (SD)** | **Mean (SD)** | |
| 6 | 49 | 57.6 (3.6) | 38 | 57.8 (2.5) | 57.7 (3.1) | 0.745 |
| 7 | 54 | 60.5 (4.9) | 38 | 59.7 (4.2) | 60.2 (4.6) | 0.382 |
| 8 | 70 | 61.6 (5.8) | 70 | 63.9 (6.3) | 62.8 (6.1) | 0.029 |
| 9 | 59 | 64.4 (7.4) | 62 | 64.4 (8.0) | 64.4 (7.7) | 0.999 |
| 10 | 62 | 65.0 (6.4) | 58 | 66.3 (6.6) | 65.6 (6.5) | 0.261 |
| 11 | 33 | 67.8 (5.5) | 34 | 65.2 (4.6) | 66.5 (5.2) | 0.042 |

N, frequency; SD, standard deviation; IQR, interquartile range; cm, centimetres; *p* derived from comparison between girls and boys using Student's t-test or Mann-Whiney U-test

### PHYSIOLOGICAL RESPONSES TO THE 6MWT

*Heart rate changes*

Overall, the mean heart rate increased significantly by 6.4 (9.5) beats/ min from baseline ($p < 0.001$; Cohen'd = -0.75), corresponding to a percentage rise (*%HRdiff)* of 7.7 % and percentage of predicted maximal heart rate of 46.6 % (Figure E1A). It then dropped by 3.4 beats/min to the 5th-minute post-walk value of 89.9 beats/min (post-hoc $p <0.001$; Cohen' d = -0.40), which was still significantly higher than the baseline value by 2.9 beats/min (post-hoc $p <0.001$; Cohen's d = -0.35).

In a 2-way ANOVA to assess the sex differences in 6MWT-induced change in HR, There was significant effect of sex on heart rate changes ($F(1, 623) = 34.6$, $p < 0.001$, $\eta^2 p = 0.05$; Fig ): compared to boys, girls had significantly higher *preHR* (mean difference = 4.2, t=4.74, $p <0.001$), *postHR* (mean difference = 5.6, t=6.37, pohm<0.001), *5mHR* (mean difference = 4.2, t=4.74, $p <0.001$) and $\%PredHR_{max}$ ($p < 0.0001$).

*Six-minute walk distance and transcutaneous oxygen saturation*

Overall, $SpO_2$ changes with 6MWT was trivial, despite showing statistical significance [F (1.9, 1244) = 4.49, $p = 0.0134$, $\eta^2 p = 0.007$). Post-hoc test showed that this significance was due to a trivial drop in the $SpO_2$ from $postSpO_2$ to $5mSpO_2$ (0.2%, $p = 0.025$, Cohen's d = 0.12). Two-way ANOVA with post-hoc test showed that there was no significant difference between boys and girls in terms of their $preSpO_2$ (mean difference = 0.1%, $p = 1.00$), $postSpO_2$ (mean difference =0.2%, p=1.0) and $5mSpO_2$ (mean difference =0.2%, p=0.43) (Figure E1B).

***Six-minute walk distance and blood pressure***

Overall, there was a significant and large change in *SBP* [$F(1.8, 1126.3) = 734.8$, $p<0.001$, Cohen's d = 0.5). The mean SBP significantly increased by 10.9 mmHg from a baseline value of 103.6 mmHg ($p<.001$, Cohen's d =-1.37), dropping to 104.4 mmHg by the 5th-minute post-walk ($p<0.001$, Cohen's d ==1.28=), but the 5th min *SBP* was still significantly but negligibly above baseline value (mean difference = -0.75, p = 0.019, Cohen's d =0.09). There was no significant difference in *SBP* between girls and boys in *preSBP* (mean difference = -0.5, p=1.0), *postSBP* (mean difference = -10.7, p=1.0) and *5mSBP* (mean difference = -0.2, p=1.0) (Figure E1C).

Change in *DBP* during the 6MWT was of moderate magnitude ($F(2,1252) = 349.2$, $p<0.001$, $\eta^2p=0.36$). Overall, the mean DBP significantly increased by 7.0 (7.2) mmHg from baseline value of 61.4 (5.4) to 68.3 (7.4) in the total sample ($p<0.001$, Cohen's d = -0.99). By the 5th minute the *DBP* had recovered to 62.7 (7.1) ($p<0.001$, Cohen's d = 0.8), which was still significantly above the baseline *DBP* (mean diff = -1.3, $p<0.001$, Cohen's d = 0.19). There was no significant sex difference in the *DBP* values at baseline (mean difference = 0.5, p=1.00), immediate post-walk (mean difference = 0.4, p=1.0) and 5th-min post-walk (mean difference = 0.2, p=1.0) (Figure E1D).

**Assumptions of 2-way ANOVA**: In all the above 2-way ANOVA tests, Mauchly's test indicated that the assumption of sphericity was violated ($W > 0.75$ $p<0.001$, epsilon = 0.94), hence Huynh-Feldt correction was adopted. Assumptions of normality and equality of variances were satisfied and there was no sex interactions in the variables.

***Six-minute walk distance and rating of perceived exertion (RPE)***

The *RPE* increased significantly from a baseline median (IQR) value of 1.0 (1.0) to post-walk value of 3.0 (3.0) (W=1939.0, $p <0.001$, $r_B = -0.96$).

***Baseline spirometric variables***

Girls had significantly higher predicted *PEF* compared to boys (p = 0.019, Cohen's d (95% CI) = 0.19 (0.03, 0.34) but there was no difference in their predicted $FEV_1$ (p = 0.182).

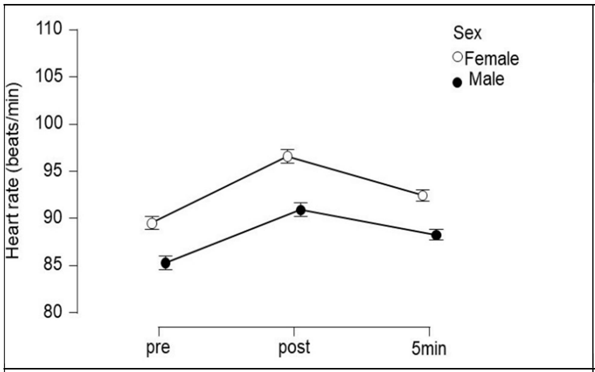


A. Heart rate changes

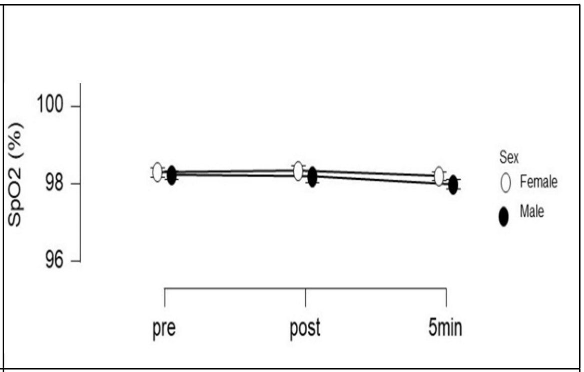


B. *$SpO_2$*

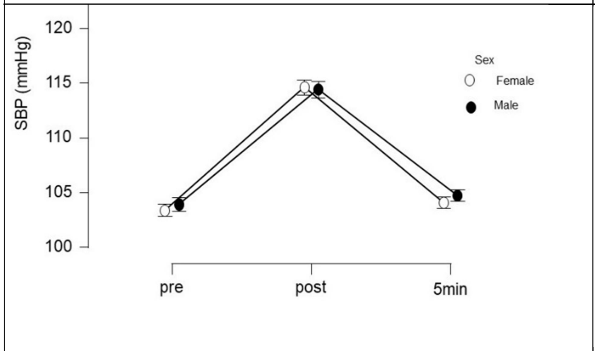


C. *SBP*

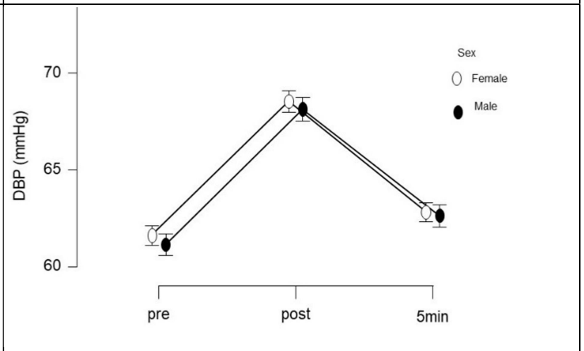


D. *DBP*

Figure E1. Changes in physiological variables (heart rate, *$SpO_2$*, *SBP* and *DBP*) during the 6MWT

*Abbreviations: $SpO_2$*, transcutaneous oxygen saturation in %; *SBP* , systolic blood pressure in mmHg

Table E3. Comparison of physiological variables between girls and boys before and after 6MWT

| Variables | Mean ± SD (Min – Max) Girls N=327 | Boys N=300 | Total N=627 | *t* | *p* |
|---|---|---|---|---|---|
| *preHR* | 89.5 ± 10.5<br>62.0 - | 85.3 ± 10.8<br>58.0 - 119.0 | 87.5 ± 10.8<br>58.0 - | 4.90 | < 0.001 |
| *postHR* | 96.6 ± 12.7<br>60.0 - | 91.0 ± 12.5<br>58.0 - 136.0 | 93.9 ± 12.9<br>58.0 - | 5.56 | <0 .001 |
| *5mHR* | 91.9 ± 10.0<br>56.0 - | 87.7 ± 10.5<br>58.0 - 118.0 | 89.9 ± 10.4<br>56.0 - | 4.70 | < 0.001 |
| Δ*HR* | 7.1 ± 9.5<br>-15 - 38 | 5.6 ± 9.5<br>-53.0 - 41.0 | 6.4 ± 9.5<br>-53.0 - 41.0 | 1.91 | 0.057 |
| *%ΔHR* | 8.3 ± 11.3<br>-14.6 - 48.7 | 7.1 ± 11.2<br>-44.5 - 50.0 | 7.7 ± 11.2<br>-44.5, 50.0 | 1.37 | 0.172 |
| *PredHR* | 201.8 ± 1.1<br>199.7 - | 201.7 ± 1.1<br>199.7 - | 201.8 ± 1.1<br>199.7 - | 1.11 | 0.268 |
| *%PredM* | 46.9 ± 6.3<br>29.9 - 68.2 | 46.1 ± 6.2<br>29.0 - 67.8 | 46.6 ± 6.4<br>28.7 - 68.7 | 5.52 | < 0.001 |
| *preSpO$_2$* | 98.3 ± 0.8<br>95 - 100 | 98.2 ± 0.8<br>96 - 100 | 98.0 ± 1.0<br>95.0 - | 1.22 | 0.222 |
| *postSpO* | 98.3 ± 1.6<br>84 - 100 | 98.2 ± 1.5<br>83 – 100 | 98.0 ± 1.0<br>88.0 - | 1.26 | 0.209 |
| 5m*SpO$_2$* | 98.2 ± 1.3<br>88 - 100 | 98.0 ± 1.4<br>87 - 100 | 98.0 ± 1.0<br>90.0 - | 2.03 | 0.043 |
| Δ$SpO_2$ | 0.04 ± 1.6<br>-14 -4 | -0.03 ± 1.6<br>-16 - 3 | 0.00 ± 1.00<br>-10.0 - 4.0 | 0.59 | 0.557 |
| *preSBP* | 103.7 ± 6.6<br>77.0-116.0 | 103.9 ± 6.5<br>76.0 - 119.0 | 103.6 ± 6.5<br>76.0- 119.0 | -1.01 | 0.311 |
| *postSBP* | 114.6 ±<br>86.0-163.0 | 114.4 ±<br>86.0 - 163.0 | 114.5 ±<br>85.0 - | 0.24 | 0.811 |
| *5mSBP* | 103.70 ±<br>75.0 - | 104.8 ± 7.8<br>75.0 - 129.0 | 104.3 ± 7.9<br>75.0 - | -1.68 | 0.094 |
| Δ*SBP* | 11.2 ± 8.8<br>-15-70 | 10.5 ± 9.1<br>-12.0 - 48.0 | 10.9 ± 8.9<br>-15.0 - 70.0 | 1.02 | 0.307 |
| *preDBP* | 61.6 ± 5.2<br>47.0 - 74.0 | 61.1 ± 5.7<br>40.0 - 75.0 | 61.4 ± 5.4<br>40.0 - 75.0 | 1.09 | 0.2761 |
| *postDBP* | 68.5 ± 7.3<br>51.0-100.0 | 68.1 ± 7.6<br>43.0 - 109.0 | 68.3 ± 7.44<br>43.0 - | 0.661 | 0.5062 |
| *5mDBP* | 62.6 ± 6.6<br>40-87 | 62.7 ± 7.6<br>41 - 96 | 62.7 ± 7.1<br>40 - 96 | -0.03 | 0.978 |
| ΔDBP | 6.9 ± 7.1 | 7.0 ± 7.3 | 7.0 ± 7.2 | -0.14 | 0.890 |

| | | | | | |
|---|---|---|---|---|---|
| | -17-42 | -14 - 37 | -17 - 42 | | |
| *preRPE* | 1.0 ± 1.0 | 1.0 ± 1.0 | 1.4 ± 1.0 | 47064* | 0.728 |
| | 1 - 5 | 1 - 6 | 1 - 6 | | |
| *postRPE* | 3.0 ± 3.0 | 3.0 ± 3.0 | 2.8 ± 3.0 | 48783* | 0.592 |
| | 1 - 7 | 1 - 10 | 1 - 10 | | |
| Δ*RPE* | 1.0 ± 2.0 | 1.0 ± 2.0 | 1.5 ± 2.0 | 48119* | 0.382 |
| | -2 - 6 | -2 - 8 | -2 - 8 | | |
| PEF | 231.5 ± | 241.8 ± | 236.4 ± | -2.47 | 0.014 |
| | 119.0 - | 121.0 - | 119.0 - | | |
| *PredPE* | 101.4 ± | 98.7 ± 13.0 | 100.1 ± | 2.35 | 0.019 |
| | 62.9-161.5 | 71.3 - 151.1 | 62.9 - | | |
| $FEV_1$ | 1.41 ± 0.34 | 1.49 ± 0.31 | 1.45 ± 0.33 | -2.93 | 0.003 |
| | 0.70 - 2.80 | 0.77 - 2.59 | 0.69 - 2.80 | | |
| *PredFE* | 98.6 ± 11.7 | 97.3 ± 12.0 | 98.0 ± 11.8 | 1.33 | 0.182 |
| | 67.4 - | 64.7- 143.8 | 64.7- 147.2 | | |

**Abbreviations**: SD, standard deviation; min, minimum; max, maximum; PreHR: pre-walk heart rate; PostHR: immediate post-walk heart rate; ΔHR: heart rate change; %ΔHR: percentage heart rate change; 5mHR: 5th-minute post-walk HR; PredHRmax:predicted maximal HR; %PredHRmax: percentage predicted maximal HR; PreSpO2: pre-walk SpO2; PostSpO2: post-walk SpO2; 5mSpO2: 5th-minute post-walk SpO2; ΔSpO2: SpO2 change; PreSBP: pre-walk SBP; PostSBP: post-walk SBP; 5mSBP: 5th-minute post-walk DBP; ΔSBP: SBP change; PreDBP: pre-walk DBP; PostDBP: post-walk DBP; 5mDBP: 5th-minute post-walk DBP; ΔDBP: DBP change; PreRPE: pre-walk rating of perceived exertion; PostRPE : post-walk rating of perceived exertion; 5mRPE: 5th-minute rating of perceived exertion; PEF: peak expiratory flow rate; %PredPEF: percentage predicted PEF; FEV1: forced expiratory volume in 1 second; %PredFEV1: percentage predicted FEV1.

Values are in mean ± SD, except otherwise stated

*Mann-Whitney U-test

MISSING VALUES: IN girls, 1 missing for PreHR, ΔHR, %ΔHR; 55 missing for 5spo2; 53 missing for 5mSBP and 5mDBP; 7 missing for preRPE; 3 missing for postRPE and 10 missing for ΔRPE. In boys, 1 missing for PreHR, ΔHR, %ΔHR and preSpO2; 39 for 5mHR, 5mSBP and 5mDBP; 3 for postSPO2; 42 for 5mSpO2; 4 for ΔSpO2, 2 for preRPE, 6 for postRPE and 8 for ΔRPE.

The presence of height at the 7th step (Table E4) destabilised the model by increasing the standard error of age from 1.47 at the preceding step (not shown) to 2.26 - 54% increase. Thus, height was manually deleted, as well as school category which is a highly variable construct to measure to yield simpler models with no violation of statistical assumptions of multivariale linear regression (Table 5).

Table E4. Exploratory stepwise Linear Regression with all significant predictors entered

| | | 95% CI | | | | | |
|---|---|---|---|---|---|---|---|
| | B | Lower | Upper | SE | *B* | *t* | *p* |
| Intercept | 253.3 | 192.2 | 314.4 | 31.10 | | 8.14 | < .001 |
| Age | 6.4 | 2.0 | 10.9 | 2.26 | 0.15 | 2.85 | 0.005 |
| Δ*HR* | 1.7 | 1.2 | 2.1 | 0.25 | 0.24 | 6.77 | < .001 |
| Δ*SBP* | 1.5 | 1.0 | 2.1 | 0.27 | 0.21 | 5.66 | < .001 |
| School category | -36.6 | -46.8 | -26.4 | 5.20 | -0.27 | -7.03 | < .001 |
| Sex | 19.6 | 10.7 | 28.4 | 4.51 | 0.15 | 4.34 | < .001 |
| Height | 1.4 | 0.7 | 2.0 | 0.35 | 0.22 | 3.90 | < .001 |

F (1,618) = 44.6, $R^2$ = 0.30, adjusted $R^2$ =0.29

All the three final models satisfied the assumptions of multiple linear regression as shown in Table E5 and Fig E3. The maximum Cook's distances were approximately 1.00 signifying absence of any influential outlier. The mean ± SD of the standardized residuals of the three equations were 0.00 ± 1.00) and ranged from -3.3 to 2.5, implying that the residuals were fairly normally distributed further confirmed by histograms and the Q-Q plots of the standardised residuals (Fig). Scatter plot of the standardised residuals with standardised predicted values shows that the residuals were evenly distributed about zero, implying homoscedasticity. There was no significant autocorrelation as the Durbin-Watson values were within 1.5 and 2.0.

Table E5. Collinearity and residual statistics for regression equations

| Equations | VIF | Cook's distance | | Residuals | | Durbin-Watson | | |
|---|---|---|---|---|---|---|---|---|
| | | Range | Mean (SD) | Range | Mean (SD) | Auto-correlation | Statistic | *p* |
| 1 | 1.1 | 0.00, 0.03 | 0.002 (0.003) | -3.3, 2.4 | 0.0002 (1.001) | 0.15 | 1.69 | <0.001 |
| 2 | 1.0 | 0.002, 0.04 | 0.002 (0.003) | -3.2, 2.5 | 0.0002 (1.001) | 0.13 | 1.74 | <0.001 |
| 3 | 1.1 | 0.00, 0.01 | 0.002 (0.002) | -2.9, 2.5 | 0.000 (1.001) | 0.13 | 1.74 | <0.001 |

Range presented as minimum, maximum values; VIF, variable inflation factor; SD, standard deviation

<table>
<tr><td>Eq 1</td><td>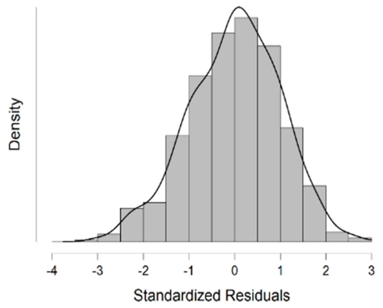
</td><td>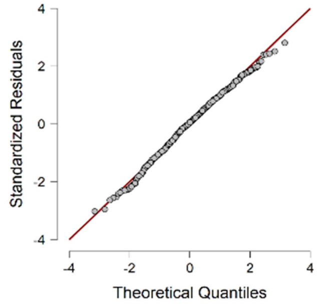
</td><td>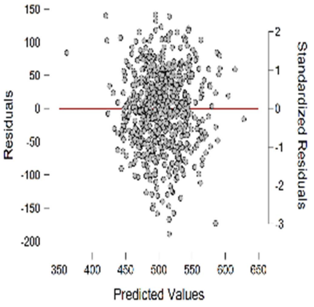
</td></tr>
<tr><td>Eq 2</td><td>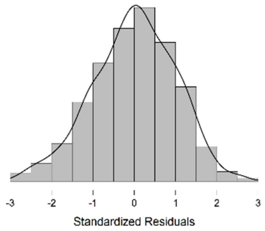
</td><td>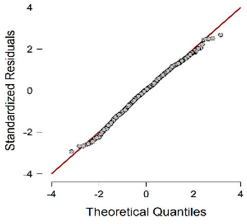
</td><td>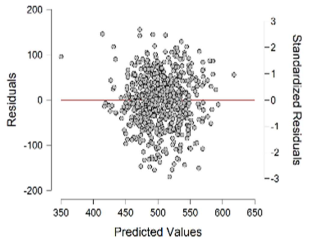
</td></tr>
<tr><td>Eq 3</td><td>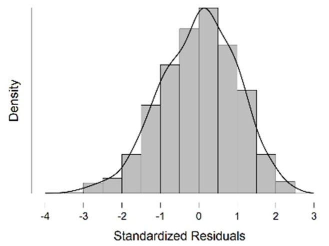
</td><td>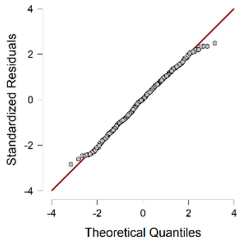
</td><td>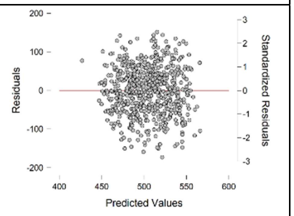
</td></tr>
<tr><td></td><td>A. Histograms of standardised residuals</td><td>B. Q-Q plots of standardised residuals</td><td>C. Scatter plots of standardised residuals and predicted values</td></tr>
<tr><td colspan="4">Fig E2. Plots of residual statistics for the 3 final equations. (A) shows that all the three histograms were bell-shaped demonstrating normally distributed residuals; (B) shows that all the three Q-Q plots were well aligned on the diagonal line; (C) shows that plots of the residuals wre essentially evenly distributed on both sides of the horizontal 0 line demonstrating homoscedasticity</td></tr>
</table>

**Table E6. Example of the clinical application of our regression equation**

An 8 year old girl with asthma walks a distance of 320 m during a 6MWT. Her pre-exercise HR and SBP are 90 bpm and 98 mmHg, respectively, while her post-exercise HR and SBP are 109 bpm and 108 mmHg respectively.

To calculate the **predicted 6MWD** using the equation

$$6MWD = 347.9 + 14Age + 1.6\Delta HR + 17.6Sex + 1.2\Delta SBP; \quad \mathit{SEE = 58.08}$$

Inserting her variables (Age =8.0 years; Sex =0 (Female = 0; Male = 1); HRdiff=post-walk HR – pre-walk HR=109-90= 19 bpm; SBPdiff = post-walk SBP – pre-walk SBP=108 – 98 = 10 mmHg) into above equation yields: 6MWD = 502.3 m

Her **percentage predicted 6MWD** is (320/502.6)x100% = 63.7% of the predicted 6MWD

To calculate her **lower limit of normal** (LLN): = predicted value – (1.645 x SEE)

= 502.3 – (1.645 x 58.08) = 406.8 m

This 8 year old girl achieved a 6MWD of 320 m which is 64% of her predicted walk distance and is lower than the lower limit of normal ($5^{th}$ percentile) value of 418.7 m expected of an 8-year old Nigerian girl (see 6MWD percentiles, Table 2). Hence, her 6MWD (functional capacity) is low and may require further evaluation.

16. Nelson SB, Lavange LM, Nie Y, Walsh JW. Questionnaires and Pocket Spirometers Provide an Alternative Approach for COPD Screening in the General Population. *Chest*

2012;142:358–366.

28. Geiger R, Strasak A, Treml B, Gasser K, Kleinsasser A, Fischer V, Geiger H, Loeckinger A, Stein JI. Six-minute walk test in children and adolescents. *J Pediatr*

2007;150:395–399.

29. WHO. WHO AnthroPlus for Personal Computers. Manual: Software for assessing growth of the world's children. *Geneva* 2009;at <http://www.who.int/growthref/tools/en/>.

30. Tanaka H, Monahan KD, Seals DR. Age-predicted maximal heart rate revisited. *J Am Coll Cardiol* 2001;37:153–156.

31. Muenchen RA. *BlueSky Statistics User's Guide*. 2020.

32. Goss-Sampson MA. *Statistical Analysis in JASP: A Students Guide v0.12*, 2nd ed. 2020. doi:10.6084/m9.figshare.9980744.

33. Andy F. Regression. *Discov Stat using SPSS*, 3rd editio. Los Angeles: SAGE Publications; 2009. p. 198–263.

34. Bland JM, Altman DG. Statistical methods for assessing agreement between two methods of clinical measurement. *Lancet* 1986;1:307–310.
